# The hyper-transmissible SARS-CoV-2 Omicron variant exhibits significant antigenic change, vaccine escape and a switch in cell entry mechanism

**DOI:** 10.1101/2022.01.03.21268111

**Authors:** Brian J. Willett, Joe Grove, Oscar A. MacLean, Craig Wilkie, Nicola Logan, Giuditta De Lorenzo, Wilhelm Furnon, Sam Scott, Maria Manali, Agnieszka Szemiel, Shirin Ashraf, Elen Vink, William T. Harvey, Chris Davis, Richard Orton, Joseph Hughes, Poppy Holland, Vanessa Silva, David Pascall, Kathryn Puxty, Ana da Silva Filipe, Gonzalo Yebra, Sharif Shaaban, Matthew T. G. Holden, Rute Maria Pinto, Rory Gunson, Kate Templeton, Pablo R. Murcia, Arvind H. Patel, The COVID-19 Genomics UK (COG-UK) Consortium, John Haughney, David L. Robertson, Massimo Palmarini, Surajit Ray, Emma C. Thomson

## Abstract

Vaccines based on the spike protein of SARS-CoV-2 are a cornerstone of the public health response to COVID-19. The emergence of hypermutated, increasingly transmissible variants of concern (VOCs) threaten this strategy. Omicron, the fifth VOC to be described, harbours 30 amino acid mutations in spike including 15 in the receptor-binding domain. Here, we demonstrate substantial evasion of neutralisation by Omicron *in vitro* using sera from vaccinated individuals. Importantly, these data are mirrored by a substantial reduction in real-world vaccine effectiveness that is partially restored by booster vaccination. We also demonstrate that Omicron does not induce cell syncytia and favours a TMPRSS2-independent endosomal entry pathway. Such marked changes in antigenicity and replicative biology may underlie the rapid global spread and altered pathogenicity of the Omicron variant.

## Main

Vaccination against SARS-CoV-2 is based primarily on vaccines that induce immunity to the spike glycoprotein. These vaccines have become the cornerstone of the global public health response to SARS-CoV-2^1^. However, their effectiveness is now being threatened by the emergence of Variants of Concern (VOC) displaying enhanced transmissibility and evasion of host immunity^2^. Of the five VOCs that have emerged, the Beta variant (B.1.351) and Gamma (P.1) variants were associated primarily with immune evasion; they spread locally but never dominated globally. In contrast, the Alpha (B.1.1.7) and Delta (B.1.617.2) VOCs spread globally and were responsible for significant waves of infections and an increase in reproduction number (R_0_). The Alpha and Delta variants harbour mutations within the polybasic cleavage site in spike (a H681 in Alpha and R681 in Delta) that enhance cleavage by furin; changes that are associated with enhanced cell entry and may contribute to increased transmissibility. While the Alpha variant spread rapidly, it was in turn replaced by the Delta variant that combined augmented transmissibility with significant immune evasion^2–5^.

Omicron (lineage B.1.1.529) is the fifth variant to be named as a VOC by the World Health Organisation (WHO) and was first detected in mid-November 2021 in Botswana, South Africa^6^ and in quarantined travellers in Hong Kong^7^. It has since split into three divergent sublineages (BA.1, BA.2 and BA.3) of which BA.1 now dominates worldwide.

Emerging data indicate that the Omicron variant evades neutralisation by sera obtained from people vaccinated with 1 or 2 doses of vaccine, especially when antibody titres are waning. Indicative studies have shown that 3 doses of spike-based vaccines may provide only partial protection from infection with this variant, including unpublished data made available as a press release from Pfizer. Immune evasion by Omicron may have contributed to the extremely high transmission rates in countries with high vaccination rates or natural immunity (R_0_ of 3-5 in the U K ^8, 9, 10, 11, 12, 13, 14, 15, 16, 17, 18.^

In this study, we investigate the antigenic and biological properties of the Omicron variant that might underlie immune evasion and increased transmission of the virus using both *in vitro* assays and real-life population data.

## Results

### Omicron displays substantial changes within spike predicted to affect antigenicity and furin cleavage

Omicron is characterised by significant changes within the RBD of the spike glycoprotein, regions targeted by class 1,2 and 3 RBD-directed antibodies, and within the N-terminal domain (NTD) supersite (Fig.1a). The G339D, N440K, S477N, T478K, Q498R and N501Y mutations enhance binding of spike to the human ACE2 receptor, while combinations such as Q498R and N501Y may enhance ACE2 binding additively^19^. Overall, the Omicron RBD binds to the human ACE2 with approximately double the affinity (x2.4) of the Wuhan RBD^8^. Deep mutational scanning (DMS) estimates at mutated sites are predictive of substantially reduced monoclonal and polyclonal antibody binding and altered binding to human ACE2 (Fig.1b)^20^. Fourteen mutations (K417N, G446S, E484A, Q493R, G496S, Q498R and to a lesser extent, G339D, S371L, S373P, N440K, S477N, T478K, N501Y and Y505H) may affect antibody binding based on a calculated escape fraction (a quantitative measure of the extent to which a mutation reduces polyclonal antibody binding by DMS). Seven Omicron RBD mutations (K417N, G446S, E484A, Q493R, G496S, Q498R and N501Y) have been shown previously to be associated with decreased antibody binding, importantly falling in epitopes corresponding to three major classes of RBD-specific neutralising antibodies (nAbs). The mutations present in spike also involve key structural epitopes targeted by several monoclonal antibodies in current clinical use. Of these, bamlanivimab, cilgavimab, casirivimab, etesevimab, imdevimab, regdanvimab and tixagevimab bind to the RBM, and neutralisation of Omicron has been shown to be negligible or absent. In contrast, sotrovimab, targets a conserved epitope common to SARS-CoV-1 and SARS-CoV-2 that is outside the RBM and has only a small reduction (x3) in neutralisation potency^21–23^. N679K and P681H mutations at the furin cleavage site (FCS) are predicted individually to increase furin cleavage, although the combination of these changes and an adjacent change (H655Y, also present in the Gamma VOC) in the vicinity of the FCS is unknown^24^.

Omicron bears three deletions (amino acids 69-70, 143-145 and 211) and an insertion (site 214) in the NTD of spike. The 69-70 deletion is also found in the Alpha and Eta (B.1.525) variants and is associated with enhanced fusogenicity and incorporation of cleaved spike into virions^25^. This 69-70 deletion is a useful proxy for estimates of Omicron prevalence in the population by S-gene target failure (SGTF) using the TaqPath^TM^ (Applied Biosystems, Pleasanton, CA) diagnostic assay. Deletions in the vicinity of amino acids 143-145 have been shown to affect a range of NTD-specific nAbs^26, 27^.

### Emergence of the Omicron variant in the UK

Despite high vaccination rates and levels of natural immunity following previous exposure in the UK, Omicron has rapidly become dominant. The evolutionary relationships of SARS-CoV-2 variants at a global level are shown in Fig.1c. The first 8 cases of Omicron were detected in the UK on the 27^th^ and 28^th^ November 2021 (2 in England and 6 in Scotland). Due to the rapid spread of Omicron, early genome sequences were highly related with an average genetic divergence between 1 and 7 single nucleotide polymorphisms (SNPs) (Fig.1d). The phylogenetic relationship to Omicron sequences from other countries was consistent with multiple introductions associated with travel to South Africa followed by community transmission.

**Fig. 1:**
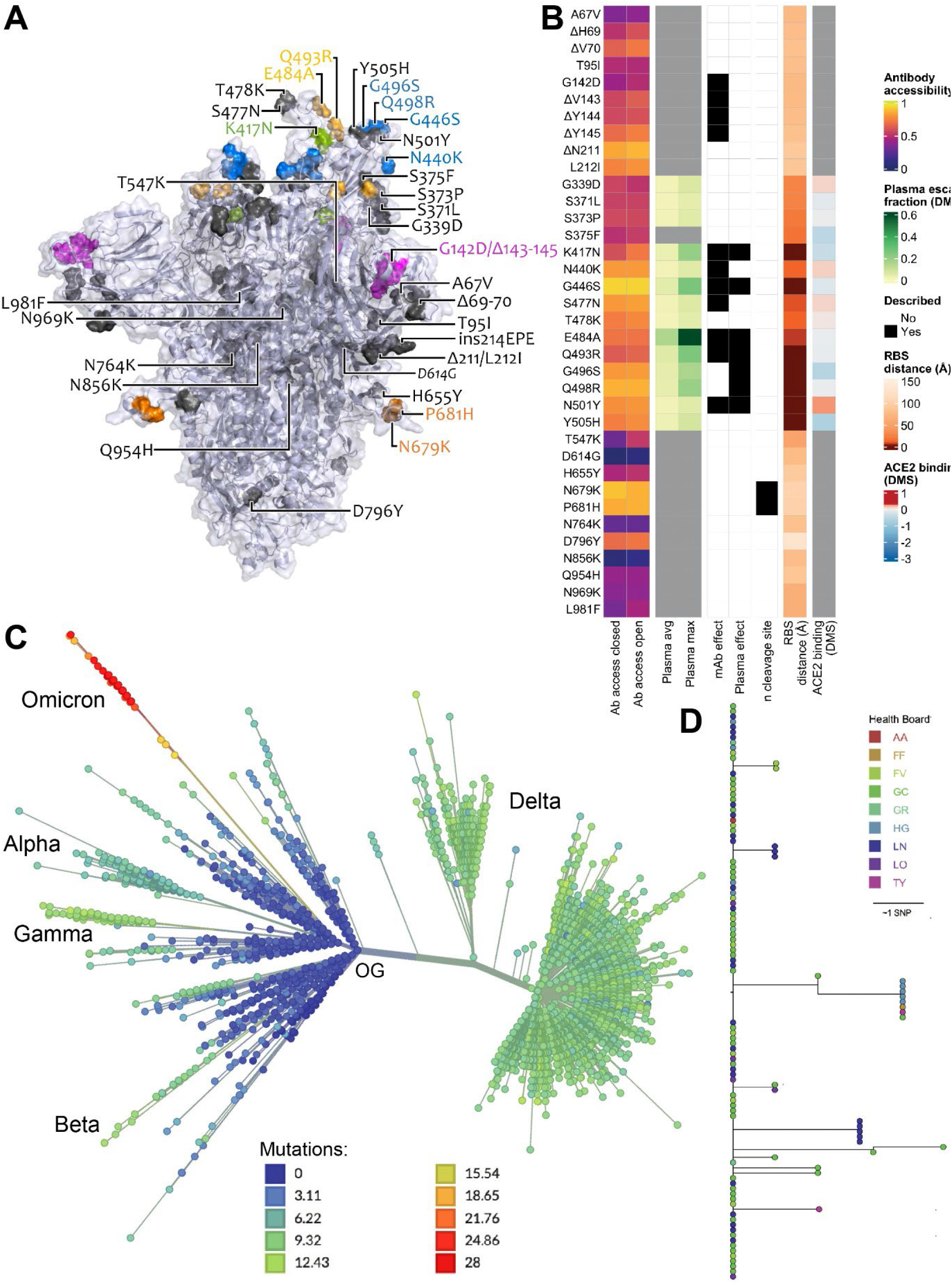
Spike amino acid changes typifying the Omicron variant. **a**, Spike homotrimer in open conformation with locations of Omicron amino acid substitutions, deletions (Δ), or insertions (ins) highlighted as spheres with opaque surface representation. Colouring highlights mutations at residues with substitutions impacting RBD-specific antibodies of classes 1 (green), 2 (yellow), and 3 (blue)^28^, or that belong to the NTD antibody supersite (magenta)^26^, or that belong to the FCS (orange), with the remainder in grey. These are annotated on the monomer with an ‘up’ receptor-binding domain. The substitution D614G which is shared by common descent by all lineage B.1 descendants is italicised. The visualisation is made using a complete spike model^29^ which is in turn based upon a partial cryo-EM structure (RCSB Protein Data Bank (PDB) ID: 6VSB^30^. **b**, Aligned heatmaps showing properties of amino acid residues or of the specific amino acid substitution present in the Omicron variant, as appropriate (insertion not shown). Structure-based epitope scores^31^ for residues in the structure of the original genotype spike in closed and open conformations are shown. For RBD residues, the results of deep mutational scanning (DMS) studies show the escape fraction (that is, a quantitative measure of the extent to which a mutation reduced polyclonal antibody binding) for each mutant averaged across plasma (‘plasma average’) and for the most sensitive plasma (‘plasma max’)^20^. Each mutation is classified as having evidence for mutations affecting neutralisation by either mAbs^27, 32–35^ or antibodies in convalescent plasma from previously infected or vaccinated individuals^20, 34–36^. Membership of the furin cleavage site is shown. The distance to ACE2-contacting residues that form the receptor-binding site (RBS) is shown (RBS defined as residues with an atom <4Å of an ACE2 atom in the structure of RBD bound to ACE2 (RCSB PDB ID: 6M0J^37^. Finally, ACE2 binding scores representing the binding constant (Δlog10 KD) relative to the wild-type reference amino acid from DMS experiments^38^. **c**, Inferred evolutionary relationships of SARS-CoV-2 from NextStrain (https://nextstrain.org/ncov/gisaid/global) with the Variants of Concern labelled. The colours of the tree tips correspond to the number of mutations causing Spike amino acid substitutions relative to the SARS-CoV-2 original genotype (OG) reference strain Wuhan-Hu-1. **d**, Inferred evolutionary relationships of the first 111 Omicron sequences in Scotland with NHS Scottish Health boards denoted: AA, Ayrshire and Arran; FF, Fife; FV, Forth Valley; GC, Great Glasgow and Clyde; GR, Grampian; HG, Highlands; LN, Lanarkshire; LO, Lothian; TY, Tayside, see key.

### Neutralising responses to Omicron (BA.1) are substantially reduced following double and partially restored following triple vaccination

Levels of nAbs in patient sera correlate strongly with protection from infection^39–42^, and reductions in neutralising activity against the Alpha and Delta variants are consistent with a reduction in vaccine effectiveness^2–5, 43^. To predict the effect of the mutations within the Omicron spike glycoprotein on vaccine effectiveness, sera collected from healthy volunteers at more than 14 days post-2^nd^ dose vaccination with either BNT162b2, ChAdOx1 or mRNA-1273 were sorted into three age-matched groups (n=24 per group, mean age 45 years). Sera were first screened by electrochemiluminescence (MSD-ECL) assay for reactivity with SARS-CoV-2 antigens (Spike, RBD, NTD or nucleoprotein (N)). The antibody responses to RBD and NTD were significantly higher (p<0.0001) in the sera from individuals vaccinated with BNT162b2 or mRNA-1273 in comparison with the ChAdOx1 vaccinees (Fig. 2a and Supplementary Table 1). In contrast, antibody responses to endemic human coronaviruses (HCoVs) (Extended data Fig. 1 and Supplementary Table 2) or influenza (Extended data Fig. 2 and Supplementary Table S3) were similar, with the exception of coronavirus OC43, where responses in BNT162b2 and ChAdOx1 vaccinees differed significantly, perhaps suggesting modulation (back-boosting) of pre-existing OC43 responses by BNT162b2 vaccination.

**Fig. 2.**
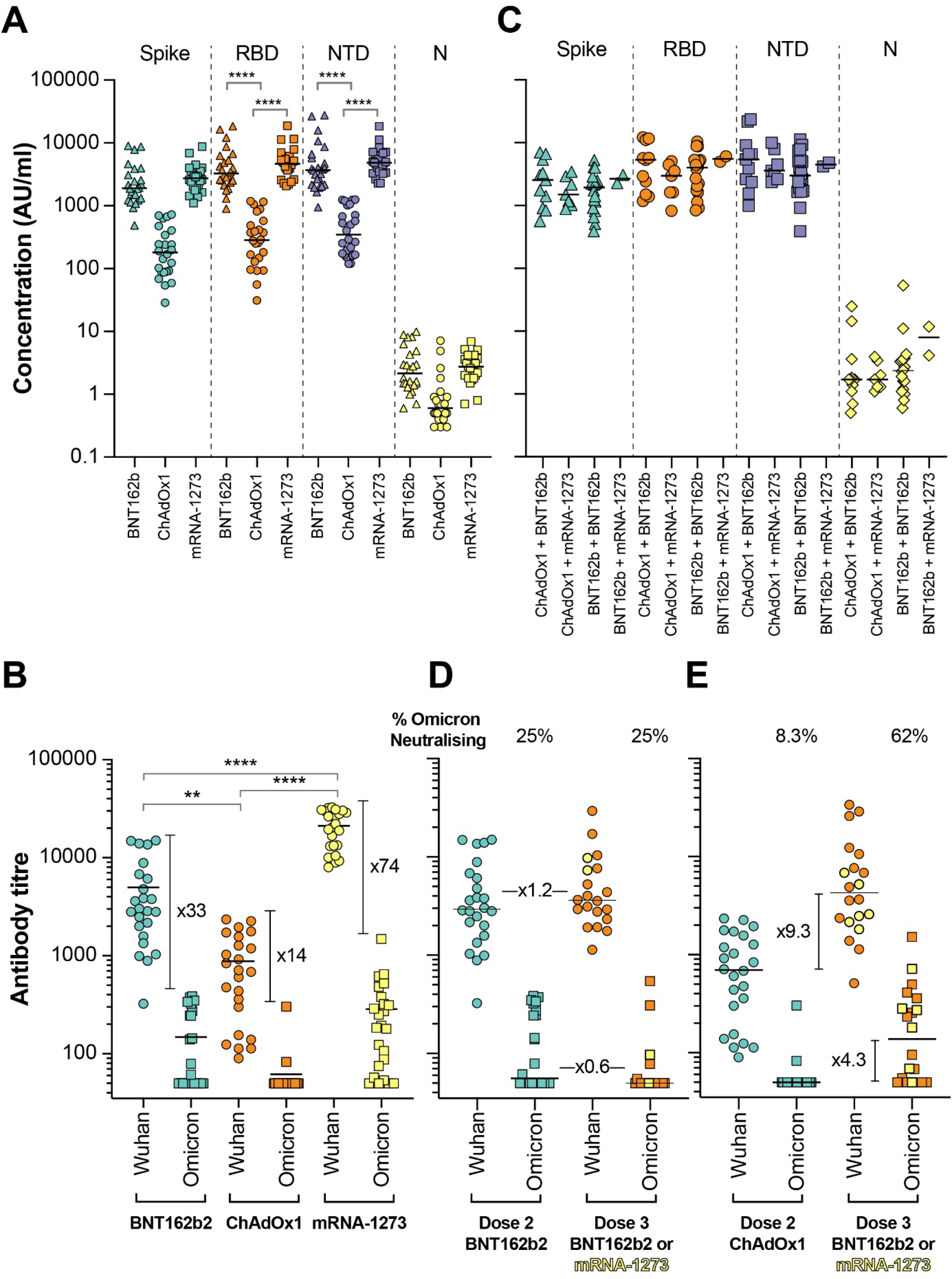
Antibody responses elicited by SARS-CoV-2 vaccination. Antibody responses were studied in three groups of individuals (n=24 per group) receiving primary vaccination with either BNT162b2, ChAdOx1 or mRNA-1273 by **a**, MSD-ECL assay or **b**, pseudotype-based neutralisation assay. **a**, Responses were measured against full-length spike glycoprotein (Spike), receptor binding domain (RBD), N-terminal domain (NTD) and nucleoprotein (N) and are expressed as arbitrary units (AU/ml). **b,** NAb responses were quantified against Wuhan or Omicron spike glycoprotein bearing HIV (SARS-CoV-2) pseudotypes. Each point represents the mean of three replicates, bar represents the group mean. To assess the effect of booster vaccines antibody responses were studied in two groups of individuals primed with two doses of either BNT162b2 or ChAdOx1 and boosted with either BNT162b2 or mRNA-1273. Reactivity against SARS-CoV-2 antigens was measured by **c,** MSD-ECL assay while neutralising activity **d,** & **e,** was measured using HIV (SARS-CoV-2) pseudotypes, as above. Green data points represent those boosted with mRNA-1273, all others received BNT162b2. In panel **d**, & **e**, % Omicron neutralising refers to the proportion of serum samples that displayed neutralising activity against Omicron pseudotypes.

Next, the nAb responses against SARS-CoV-2 pseudotypes expressing the spike glycoprotein from either Wuhan-Hu-1, or Omicron (BA.1) were compared (Fig. 2b). Vaccination with mRNA-1273 elicited the highest nAb titres (mean titre Wuhan=21,118, Omicron=285), in comparison with those elicited by vaccination with either BNT162b2 (Wuhan=4978, Omicron=148.3) or ChAdOx1 (Wuhan=882.3, Omicron=61.9). Neutralising antibody titres against Wuhan differed significantly between the three study groups. Activity against Omicron was markedly reduced in comparison with Wuhan, reduced by 33-fold for BNT162b2, 14-fold for ChAdOX1 and 74-fold for mRNA-1273 (Supplementary Table 4). While the fold change in neutralisation was lowest in recipients of the ChAdOx1 vaccine and highest in recipients of the mRNA-1273 vaccine, absolute neutralisation values were highest in mRNA-1273 followed by BNT162b2 and ChAdOx1. Neutralisation was lowest in the ChAdOx1 group, however it is important to note that this was given to older patients during early vaccine rollout in the UK, especially to vulnerable patients in nursing homes and was not recommended in young adults less than 40 years of age.

Next, samples were analysed from vaccine recipients at least 14 days post booster vaccination (third dose). Participants had been primed with two doses of either ChAdOx1 or BNT162b2, followed by a third dose of either BNT162b2 (full dose) or mRNA-1273 (half dose; 50µg). All sera reacted strongly with SARS-CoV-2 antigens by MSD-ECL, with no significant differences between the four groups (Fig. 2c and Supplementary Table 5). Antibody responses to HCoVs (Extended data Fig. 3 and Supplementary Table 6) or influenza (Extended data Fig. 4 and Supplementary Table 7) were similar, with the exception of influenza Michigan H1, where responses in ChAdOx1-primed and BNT162b2 or mRNA-1273-boosted groups differed significantly, likely reflecting co-administration of influenza booster vaccines during the booster campaign. Two vaccine recipients boosted with BNT162b2 displayed weak reactivity with nucleocapsid (Fig. 2c), suggesting previously undetected exposure to SARS-CoV-2. Sera from vaccine recipients primed with BNT162b2 and boosted with either BNT162b2 or mRNA-1273 displayed similar titres of nAb against Wuhan to the samples collected post-dose 2 (Fig. 2d). In contrast, vaccination of individuals primed with ChAdOx1 with a booster dose of either BNT162b2 or mRNA-1273 resulted in a marked increase in antibody titre (9.3-fold increase) against Wuhan relative to the low titres after dose 2 (Fig. 2e and Supplementary Table 8). The marked increase in antibody titre in ChAdOx1-primed individuals (Extended data Fig. 5) emphasises the importance of the third dose booster in this population. Indeed, following boost with either BNT162b2 or mRNA-1273, anti-Wuhan nAb titres in the ChAdOx1-primed group were not significantly different from those primed with BNT162b2 (Supplementary Table 8). NAb titres against Omicron were lower in both booster study groups and did not differ significantly in titre (Supplementary Table 8). However, absolute numbers displaying measurable Omicron neutralising activity were higher in the ChAdOx1-primed group (13/21, 62%) compared with the BNT162p2 primed group (5/20, 25%) (Fig. 2d, Fig. 2e).

### Vaccine effectiveness against the Omicron variant is reduced compared to the Delta variant

A logistic additive model with a test negative case control design was used to estimate relative vaccine effectiveness against becoming a confirmed case with Delta (4911 cases) and/or Omicron (6166 cases) in a population of 1.2 million people in the largest health board in Scotland, NHS GG&C, between 22^nd^ and 28^th^ December 2021. Demographic data are shown in Supplementary Table 9. The timing of first doses of vaccination are shown in Fig.3a and the occurrence of sequenced/confirmed infections with different variants in vaccine recipients over time is shown in Fig.3b. Infection status for Omicron and Delta was modelled by number and product type of vaccine doses, previous infection status, sex, SIMD quartile, and age (to control for demographic bias). Immunosuppressed individuals were removed from the analysis to ensure case-positivity could be attributed to vaccine escape rather than an inability to mount a vaccine response, with immunosuppression status derived from a list of those in GG&C shielding due to immunosuppression or listed as being given immunosuppressant drugs. Age and time since vaccination were each modelled as single smooth effects using thin plate regression splines^44^.

**Fig. 3.**
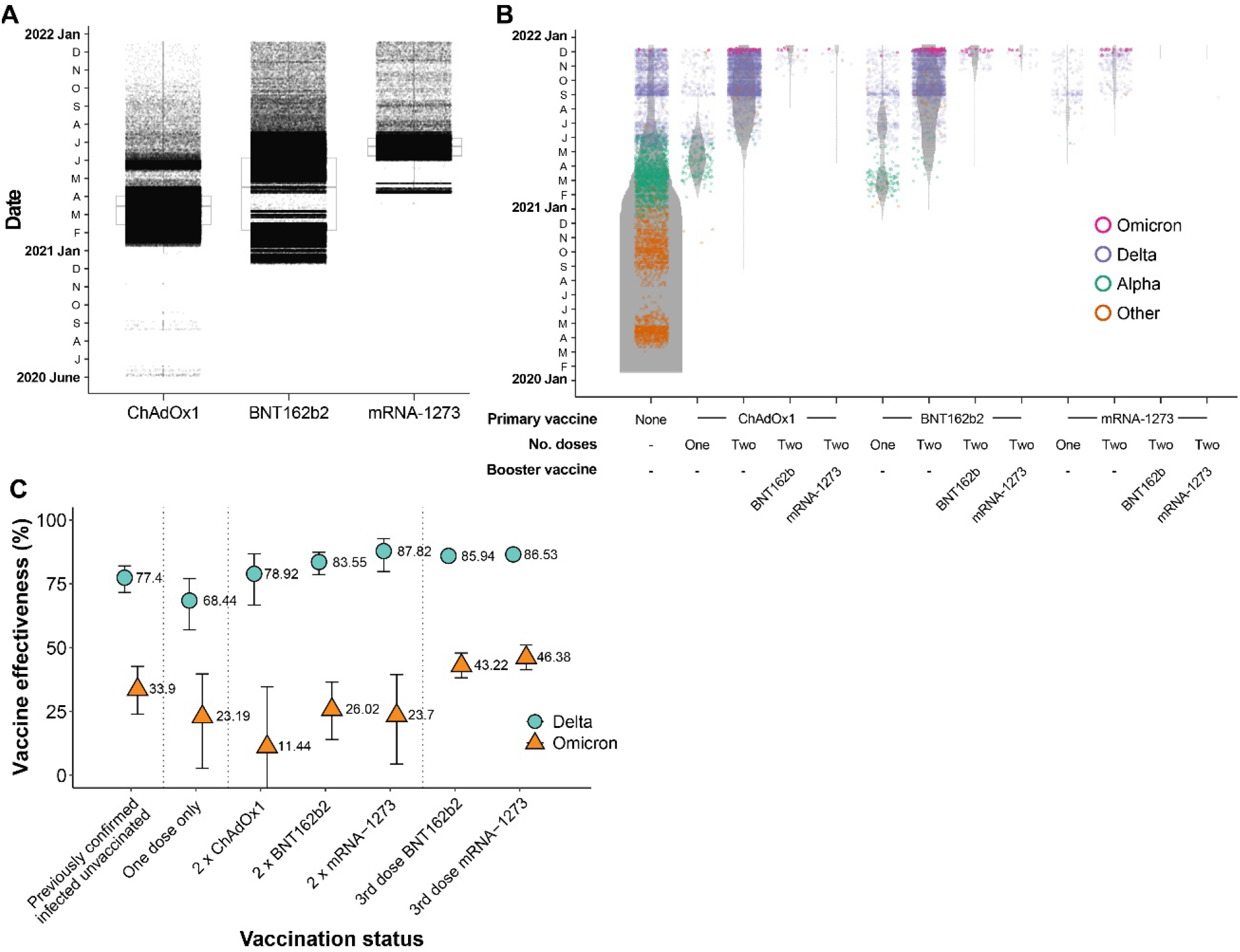
Vaccine deployment and vaccine effectiveness estimates. **a**, Denominator (violin) plot showing populations of test positive and test negative cohorts in NHS GG&C, with the widths of the grey bands represent the populations in each group at each time point. VOC classification of sequenced cases are overlaid as a dotplot, with points coloured by their VOC and a random jitter applied along the x-axis for visual clarity. **b,** Boxplots of date of first administered vaccine dose by vaccine product for the population of NHS Greater Glasgow and Clyde (NHS GG&C) aged 18 and older. The box limits are the quartiles and the centre line is the median, with whisker length of 1.5 times the interquartile range. Outliers are shown as dots outwith the whisker range. Data points are overlaid as a dotplot with points shown as black dots, with a random jitter along the x-axis applied for visual clarity. **c,** Error bar plot of estimated vaccine effectiveness against testing positive for Delta and Omicron SARS-CoV-2 infection in the population of over 18s in NHS GG&C who were tested between 6^th^ and 12^th^ December 2021. The points and corresponding text represent the estimated vaccine effectiveness (%) for each group, for each variant, with the error bar endpoints representing the endpoints of the corresponding 95% CIs. The additive effect of infection-acquired immunity was calculated for the entire population and plotted for the unvaccinated cohort.

The protection from vaccine-acquired and infection-acquired immunity were estimated as being markedly reduced against Omicron compared with Delta. Estimates of vaccine effectiveness in recent recipients (at 14 days post-dose) were only 11.44% for full primary courses of ChAdOx1 against Omicron and 78.92% against Delta. For two doses of mRNA vaccines, vaccine effectiveness was significantly lower for Omicron than Delta; BNT162b2 (26.02% versus 83.55%) and mRNA-1273 (23.70% versus 87.82%) (Fig.3c). These responses were similar following a third booster dose of BNT162b2 or mRNA-1273 against Delta (85.94% and 86.53%, respectively), but increased significantly against Omicron (43.22% and 46.38%, respectively). These estimates are in keeping with those reported recently against symptomatic infection in England where vaccine effectiveness was estimated as 71.4% and 75.5% for ChAdOx1 and BNT162b2 primary course recipients boosted with BNT162b2, respectively^9^.

Next, we estimated the additive protective effect of previous natural infection. Infection-acquired immunity directed against other VOCs may be broader in nature and may wane more slowly than that induced by vaccines^45–47^. The level of protection following previous infection was 33.9% for Omicron, and 77.4% for Delta. This level of protection was greater than that observed following two doses of vaccine for Omicron but did not reach the levels attained by those who had never had natural infection and had received third dose boosters for either Omicron or Delta. Collectively, these results emphasise the importance of booster vaccines. Importantly, vaccine-mediated protection against severe disease is likely to be more durable than that against detected infection^48^.

### Absence of syncytia in Omicron-infected cells

Our data demonstrate that antigenic change in Omicron permits evasion of vaccine induced immunity, however, the constellation of spike mutations in Omicron suggest that functional change may also contribute to its rapid transmission (Fig.1a). Therefore, we investigated the virological properties of live Omicron isolated from a patient sample. SARS-CoV-2 particles can achieve membrane fusion at the cell surface following proteolytic activation of spike by the plasma membrane protease TMPRSS2. This property also permits spike-mediated fusion of SARS-CoV-2 infected cells with adjacent cells resulting in syncytia^49^; a feature that has been associated with severe disease^50^. Moreover, the Delta variant has been shown to exhibit enhanced fusion compared to the Alpha and Beta variants^51^.

A split GFP cell-cell fusion system^52^ was used to quantify syncytia formation n by Omicron, Delta and first wave Wuhan D614G virus (Fig. 4a). Cells expressing split GFP were infected with Wuhan-D614G, Delta or Omicron and the levels of the reconstituted GFP signal following cell-cell fusion was determined in real time (Fig. 4c). In addition, infected cells were probed by indirect immunofluorescence assay to assess viral replication by the detection of the viral nucleocapsid protein (Fig. 4b). The Delta variant exhibited the highest levels of cell fusion followed by Wuhan D614G. In contrast, the Omicron variant failed to form syncytia. This failure was not due to lack of infection as immunofluorescent detection of nucleocapsid protein confirmed viral replication by Omicron, Wuhan D614G and Delta^18^.

**Fig. 4.**
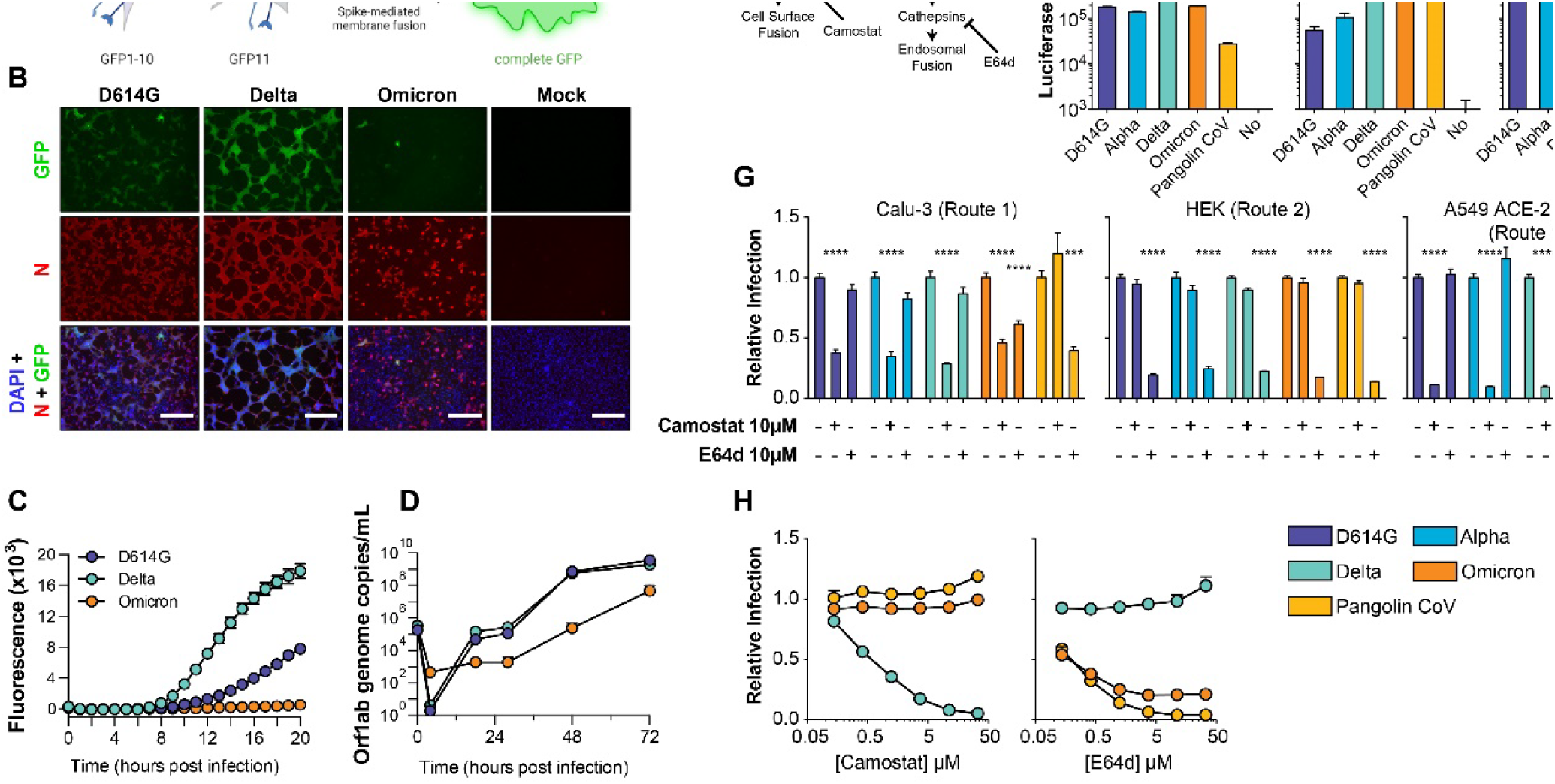
Omicron exhibits reduced syncytia formation and has switched entry route preference. **a**, Schematic representation of the split GFP system, used in this study to quantify virus induced cell fusion. This system is based on co-culture of two different cell lines (GFP-10 and GFP-11) expressing split GFP molecules. Upon virus-induced cell fusion, the intact GFP molecule is reconstituted, and the resulting signal can be detected and quantified. **b,** GFP-10 and GFP-11 A549 ACE2 TMPRSS2 cells were co-cultured and infected with Wuhan D614G, Delta and Omicron and incubated in a CLARIOStar Plus (BMG LABTECH) at 37°C / 5% CO_2_. Micrographs display cells 22 hours post infection: reconstituted GFP (green), N viral nucleocapsid (red), and DAPI nuclear counter stain (blue). Scale bars 100µm. **c,** To quantify, GFP signal was measured every 30 min for 20h. Omicron infected cells showed only background levels of GFP signal. **d,** Calu-3 cells were infected with Wuhan D614G, Delta and Omicron and supernatants were collected at the indicated times and assessed by RT-qPCR. Omicron display reduced replication kinetics compared to Wuhan D614G and Omicron. **e,** SARS CoV-2 entry can occur via two routes. Route 1 permits rapid fusion at the cell surface following proteolytic processing by TMPRSS2. In Route 2 fusion occurs following endocytosis after processing by cathepsin B or L. Route 1 and 2 can be specifically inhibited using the protease inhibitors Camostat and E64d, respectively. **f,** SARS-CoV-2 pseudotype infection of the stated cell lines, data represent mean luciferase values from one representative experiment. In Calu-3 cells Route 1 entry predominates whereas HEK exclusively support Route 2, A549 ACE2 TMPRSS2 cells permit both routes. Pangolin CoV spike is included as a control; it can only achieve entry via Route 2. Pseudotypes without viral glycoproteins (No) are included as a negative control. **g,** Relative SARS-CoV-2 pseudotype infection (compared to untreated control) of cells treated with 10μM protease inhibitors. Data represent mean of four replicates, error bars indicate standard error of the mean, asterisks indicate statistical significance (ANOVA). **h,** Titration of Camostat and E64d against Delta, Omicron and Pangolin CoV in A549 ACE2 TMPRSS2 cells, data points represent mean relative infection, compared to untreated control.

### Reduced replication kinetics of Omicron in lung epithelial cells

The replication of Omicron, Delta and Wuhan D614G was compared in Calu-3, a human lung epithelial cell line. Wuhan D614G and Delta displayed comparable replication kinetics over a period of 72 hours, with visible CPE between 48-72hpi (Fig. 4d). In contrast, the titres of Omicron were at least an order of magnitude lower at each time point compared to Wuhan D614G and Delta. These observations are consistent with attenuated replication of Omicron in lower respiratory tissues as recently reported^18, 53^.

### Omicron spike has switched entry route preference

Entry of SARS-CoV-2, and related coronaviruses, can proceed via two routes^54^. Cell surface fusion following proteolysis by TMPRSS2, as described above (“Route 1”, Fig. 4e), or fusion from the endosome after endocytosis and activation by the endosomal proteases Cathepsin B or L (“Route 2”, Fig. 4e). The ability of SARS-CoV-2 to achieve cell surface fusion is dependent on its S1/S2 polybasic cleavage site; this is absent from most closely related sarbecoviruses, which are confined to endosomal fusion^55–57^. Given the reduced fusogenicity and replication kinetics of Omicron, HIV pseudotypes were used to evaluate entry route preference. We tested Wuhan D614G, Alpha, Delta and Omicron spike, while Pangolin CoV (Guangdong isolate) spike was included as a control. Pangolin CoV spike exhibits high affinity interactions with human ACE2 but lacks a polybasic cleavage site and, therefore, enters via the endosome only^58–61^.

Calu-3 cells support cell surface (Route 1) fusion predominantly, owing to their high endogenous expression of TMPRSS2^56, 62^; in these cells, Delta yielded the highest infection, being ∼4 fold higher than Omicron (Fig. 4f). Pangolin CoV infection was low, indicating that Calu-3 cells do not support robust endosomal entry. In contrast, HEK only support endosomal entry and in these cells Pangolin CoV had high infection. Notably, Omicron also achieved high infection in HEK cells, producing ∼10-fold greater signal than Delta. This suggests that Omicron, like Pangolin CoV, is optimised for endosomal entry. All pseudotypes exhibited robust infection in A549-ACE2-TMPRSS2, where both entry routes are available^63, 64^.

Entry pathway preference was further investigated using protease inhibitors targeting either TMPRSS2 (Camostat) or cathepsins (E64d)^55^. In Calu-3 cells, all SARS-CoV-2 pseudotypes were inhibited by Camostat, whereas only Omicron exhibited E64d sensitivity, indicating that a component of infection occurs via endosomal entry (Fig. 4g). In HEK cells, all pseudotypes were inhibited by E64d, whereas Camostat was non-inhibitory; this confirms that only endosomal entry is available in these cells. Inhibitor treatment in A549 ACE2 TMPRSS2 provided the clearest evidence of altered entry by Omicron. D614G, Alpha and Delta were potently inhibited by Camostat, but not E64d. For Omicron, and Pangolin CoV, this pattern was reversed, suggesting a binary switch from cell surface to endosomal fusion; this conclusion was supported by titration of either inhibitor in A549 ACE2 TMPRSS2 cells (Fig. 4h).

These data indicate that, whilst Delta is optimised for fusion at the cell surface, Omicron preferentially achieves entry through endosomal fusion; this biological about-face may impact transmission, cellular tropism and pathogenesis. Moreover, this switch away from TMPRSS2-mediated activation offers a mechanistic explanation for reduced syncytia formation by Omicron infected cells.

## Discussion

The Omicron variant represents a major change in biological function and antigenicity of SARS-CoV-2. In this study, we demonstrate substantial immune escape of the Omicron variant. We present clear evidence of vaccine failure in dual-vaccinated individuals and partial restoration of immunity following a third booster dose of mRNA vaccine. In addition, we demonstrate a shift in the SARS-CoV-2 entry pathway from cell surface fusion, triggered by TMPRSS2, to cathepsin-dependent fusion within the endosome. This fundamental biological shift may affect the pathogenesis and severity of disease and requires further evaluation in population-based studies.

Using sera from double vaccine recipients, Omicron was found to be associated with a drop in neutralisation greater in magnitude than that reported in all other variants of concern (including Beta and Delta). Boosting enhanced neutralising responses to both the vaccine strain (Wuhan) and Omicron, particularly in recipients of ChAdOx1, but did not completely overcome the inherent immune escape properties of Omicron. Importantly, we did not assess the impact of vaccination on clinical severity of disease which is likely to be much higher than detection of infection. Protection against severe disease is longer lasting than prevention of infection. We also did not measure the impact of vaccination on T cell immunity which may be better preserved as only 14% of CD8+ and 28% of CD4+ epitopes are predicted to be affected by key Omicron mutations^12^.

The probability of infection with Omicron versus infection with the preceding Delta variant was significantly higher in double vaccine recipients, in keeping with the neutralisation data. A third dose of mRNA vaccine substantially reduced the probability of infection but did not restore immunity fully.

The emergence of a highly transmissible variant that is associated with escape from vaccine-induced immune responses means that over time, Omicron-specific vaccines may be required if disease severity was high, either directed at the general population or vulnerable groups. Early indications in young people are that Omicron infection is 40-70% less severe than Delta infection^65, 66^ similar calculations in the most vulnerable part of the population over the age of 40 years are awaited.

Genotypic changes in new variants have previously been shown to alter viral phenotype by modulating innate immune responses as well as evasion of the adaptive immune response^67, 68^. Additionally, mutations can alter spike functionality to impact transmission and pathogenesis^24^. Such changes may have provided emergent viruses with a selective advantage in lung cells and primary human airway epithelial cells. Enhanced spike activation by the plasma membrane protease TMPRSS2, may have enabled more rapid cell surface fusion^54^. In this study, we found that the Omicron variant has switched entry pathway to use TMPRSS2-independent endosomal fusion preferentially, a major change in the biological behaviour of the virus. This switching in the mechanism of fusion activation also manifests in reduced syncytia formation in infected cells, likely to reduce the cell-to-cell transmission characteristics of other variants. These properties have the potential to change cellular tropism and disease pathogenesis significantly. However, even a variant that is less virulent with a very high transmission rate may continue to present a substantial risk to older people and those with co-morbidities, especially those with immunosuppression. Moreover, our work demonstrates that SARS-CoV-2 exhibits high antigenic and functional plasticity; further fundamental shifts in transmission and disease should be anticipated.

## Methods

### Cells

Calu-3 are human lung adenocarcinoma epithelial cells. Caco-2 are an immortalized cell line derived from human colorectal adenocarcinoma, primarily used as a model of the intestinal epithelial barrier. A549 cells, a human alveolar adenocarcinoma line, were modified to stably express human ACE-2 and TMPRSS2. Human embryonic kidney (HEK293T) cells were used in pseudotype production. Baby Hamster Kidney clone 21 cells and Vero ACE-2 TMPRSS2 cells were used in the isolation of live Omicron SARS-CoV-2. All cell lines were maintained at 37°C and 5% CO_2_ in DMEM supplemented with 10% foetal bovine serum (FBS), except for Calu-3 cells which were supplemented with 20% FBS.

### Generation of cell line expressing human ACE2 receptor

Lentiviral vectors encoding human *ACE2* (GenBank NM_001371415.1) were produced as described previously^69^ and BHK-21 transduced cells were selected with 200µg/ml of hygromycin B.

### Generation of cell lines used for fusion assays

Retrovirus vectors were produced by transfecting HEK-293T cells with plasmid pQCXIP-GFP1-10 (Addgene #68715) or pQCXIP-BSR-GFP11 (Addgene #68716)^52^ and packaging vectors expressing MLV gal-pol and VSV-G using Lipofectamine 3000 (Invitrogen) according to manufacturer’s instructions. Cell supernatants were harvested 24-48h post-transfection, pooled, clarified by centrifugation and filtered. One mL of each supernatant was used to transduce A549-Ace2-TMPRSS2 (AAT) cells^69^ in presence of Polybrene (Merck). Two days post-transduction, the supernatant was replaced with selection medium (DMEM 10% FBS 1µg/mL puromycin) and cells incubated until complete death of the untrasduced control cells were observed. The resulting puromycin-resistant cells (termed AAT-GFP1-10 and AAT-BSR-GFP11) were used in fusion assays.

### Virus isolation from clinical samples

Nasopharyngeal swabs of patients infected with Omicron were collected with biorepository ethical approval (reference 20/ES/0061) in virus transport medium and resuspended in serum-free DMEM supplemented with 10 µg/ml gentamicin, 100 units/ml penicillin-streptomycin and 2.5µg/ml amphotericin B to a final volume of 1.5ml. Virus isolation was attempted in BHK-21 cells stably expressing the human ACE2 protein (BHK-hACE2) and VERO cells stably expressing ACE2 and TMPRSS2 (VAT^69^). The infected cells were incubated at 37°C and monitored for signs of cytopathic effect (CPE) and the presence of viral progeny in the medium by RT-qPCR. While no CPE was observed in any of the infected cells, RT-qPCR at 5 days post-infection (dpi) confirmed the presence of the virus derived from two of the five samples (referred to hereafter as 204 and 205) in the medium of BHK-hACE2, but not VAT cells (Extended data Fig. 7a). An aliquot of the clarified medium containing approximately 4×10^4^ viral genomes of the P0 stocks of samples 204 and 205 was used to infect VAT, BHK-ACE2 and Calu-3 cells. No CPE was observed in the infected cells but once again, virus replication was confirmed in BHK-hACE2 and Calu-3 by RT-qPCR. Supernatants (termed P1) from infected Calu-3 cells at 3 dpi were collected and virus titrated by both focus forming assay and RT-qPCR. The virus reached more than 100-fold higher titres in Calu-3 cells compared to BHK-hACE2 (Extended data Fig. 7b). Further passage of sample 205-derived P1 virus in both Calu-3 and Caco-2 yielded equivalent genome copy numbers in both cell lines (Extended data Fig. 7b). CPE was observed at 3 dpi in both Calu-3 and Caco-2 cells (not shown). The medium (termed P2) of infected Calu-3 and Caco-2 cells was collected at 4 dpi, titrated and used in subsequent experiments.

### Measurement of SARS-CoV-2, HCoVs and influenza antibody response by electrochemiluminescence

IgG antibody titres were measured quantitatively against SARS-CoV-2 trimeric spike (S) protein, N-terminal domain (NTD), receptor binding domain (RBD) or nucleocapsid (N), human seasonal coronaviruses (HCoVs) 229E, OC43, NL63 and HKU1; and influenza A (Michigan H1, Hong Kong H3 and Shanghai H7) and B (Phuket HA and Brisbane) using MSD V-PLEX COVID-19 Coronavirus Panel 2 (K15369) and Respiratory Panel 1 (K15365) kits. Multiplex Meso Scale Discovery electrochemiluminescence (MSD-ECL) assays were performed according to manufacturer instructions. Briefly, 96-well plates were blocked for one hour. Plates were then washed, samples were diluted 1:5000 in diluent and added to the plates along with serially diluted reference standard (calibrator) and serology controls 1.1, 1.2 and 1.3. After incubation, plates were washed and SULFO-TAG detection antibody added. Plates were washed and were immediately read using a MESO Sector S 600 plate reader. Data were generated by Methodological Mind software and analysed using MSD Discovery Workbench (v4.0). Results are expressed as MSD arbitrary units per ml (AU/ml). Reference plasma samples yielded the following values: negative pool - spike 56.6 AU/ml, NTD 119.4 AU/ml, RBD 110.5 AU/ml and nucleocapsid 20.7 AU/ml; SARS-CoV-2 positive pool - spike 1331.1 AU/ml, NTD 1545.2 AU/ml, RBD 1156.4 AU/ml and nucleocapsid 1549.0 AU/ml; NIBSC 20/130 reference - spike 547.7 AU/ml, NTD 538.8 AU/ml, RBD 536.9 AU/ml and nucleocapsid 1840.2 AU/ml.

### Measurement of virus neutralising antibodies using viral pseudotypes

Pseudotype-based neutralisation assays were carried out as described previously ^70 71 72^. Briefly, HEK293, HEK293T, and 293-ACE2 ^71^ cells were maintained in Dulbecco’s modified Eagle’s medium (DMEM) supplemented with 10% FBS, 200mM L-glutamine, 100µg/ml streptomycin and 100 IU/ml penicillin. HEK293T cells were transfected with the appropriate SARS-CoV-2 S gene expression vector (wild type or variant) in conjunction with p8.91 ^73^ and pCSFLW ^74^ using polyethylenimine (PEI, Polysciences, Warrington, USA). HIV (SARS-CoV-2) pseudotypes containing supernatants were harvested 48 hours post-transfection, aliquoted and frozen at -80°C prior to use. S gene constructs bearing the WUHAN (D614G) and Omicron (B.1.1.529) S genes were based on the codon-optimised spike sequence of SARS-CoV-2 and generated by GeneArt (ThermoFisher). Constructs bore the following mutations relative to the Wuhan-Hu-1 sequence (GenBank: MN908947): WUHAN (D614G) – D614G; Omicron (BA.1, B.1.1.529) - A67V, Δ69-70, T95I, G142D/Δ143-145, Δ211/L212I, ins214EPE, G339D, S371L, S373P, S375F, K417N, N440K, G446S, S477N, T478K, E484A, Q493R, G496S, Q498R, N501Y, Y505H, T547K, D614G, H655Y, N679K, P681H, N764K, D796Y, N856K, Q954H, N969K, L981F. 293-ACE2 target cells were maintained in complete DMEM supplemented with 2µg/ml puromycin.

Neutralising activity in each sample was measured by a serial dilution approach. Each sample was serially diluted in triplicate from 1:50 to 1:36450 in complete DMEM prior to incubation with HIV (SARS-CoV-2) pseudotypes, incubated for 1 hour, and plated onto 239-ACE2 target cells. After 48-72 hours, luciferase activity was quantified by the addition of Steadylite Plus chemiluminescence substrate and analysis on a Perkin Elmer EnSight multimode plate reader (Perkin Elmer, Beaconsfield, UK). Antibody titre was then estimated by interpolating the point at which infectivity had been reduced to 50% of the value for the no serum control samples.

### Protease inhibitor studies

To selectively inhibit either cell surface or endosomal fusion of SARS-CoV-2, cells were pre-treated for one hour with 10µM of either Camostat mesylate (referred to hence forth as Camostat) or E64d prior to inoculation with pseudotype. In these studies, spike proteins from Alpha and Delta VOCs, and Guangdong isolate Pangolin coronavirus (GISAID ref EPI_ISL_410721) were used as controls.

### Viral RNA extraction and RT-qPCR

Viral RNA was extracted from culture supernatants using the RNAdvance Blood kit (Beckman Coulter Life Sciences) following the manufacturer’s recommendations. RNA was used as template to detect and quantify viral genomes by duplex RT-qPCR using a Luna® Universal Probe One-Step RT-qPCR Kit (New England Biolabs, E3006E). SARS-CoV-2 specific RNAs were detected by targeting the N1 gene from the CDC panel as part of the SARS-CoV-2 Research Use Only qPCR Probe Kit (Integrated DNA Technologies) and the ORF1ab gene using the following set of primers and probes: SARS-CoV-2_Orf1ab_Forward 5’ GACATAGAAGTTACTGG&CGATAG 3’, SARS-CoV-2_Orf1ab_Reverse 5’ TTAATATGACGCGCACTACAG 3’, SARS-CoV-2_Orf1ab_Probe ACCCCGTGACCTTGGTGCTTGT with HEX/ZEN/3IABkFQ modifications. SARS-CoV-2 RNA was used to generate a standard curve and viral genomes were quantified and expressed as number of Orf1ab RNA molecules /ml of supernatant. All runs were performed on the ABI7500 Fast instrument and results analysed with the 7500 Software v2.3 (Applied Biosystems, Life Technologies).

### Genome Sequencing and analysis

Sequencing was carried out by the UK public health agencies (UKHSA/PHE, PHS, PHW and PHNI) and by members of the COG-UK consortium using the ARTIC protocol as previously described. Sequences were aligned by mapping to the SARS-CoV-2 reference Wuhan-Hu-1 using Minimap2 ^75^. Prior to phylogenetic analysis 85 sites exhibiting high genetic variability due to data quality issues in overseas sequencing labs were excluded using a masking script in Phylopipe (https://github.com/cov-ert/phylopipe). The phylogenetic tree was constructed with the maximum likelihood method FastTree2 ^76^ using a JC+CAT nucleotide substitution model.

### Replication curve

Calu-3 cells were seeded in a 96-well plate at a cell density of 3.5×10^4 cells per well. Cells were infected with the indicated viruses using the equivalent of 2×10^4 Orf1ab genome copies/well in serum-free RPMI-1640 medium (Gibco). After one hour of incubation at 37°C, cells were washed three times and left in 20% FBS RPMI-1640 medium. Supernatants were collected at different times post-infection and viral RNA extracted and quantified as described above.

### Fusion assay

AAT-GFP1-10 and AAT-BSR-GFP11 cells were trypsinized and mixed at a ratio of 1:1 to seed a total of 2×10^4 cells/well in black 96-well plate (Greiner) in FluoroBrite DMEM medium (Thermo Fischer Scientific) supplemented with 2% FBS. Next day, cells were infected with the indicated viruses using the equivalent of 10^6 Orf1a genome copies/well in FluoroBrite DMEM 2% FBS. GFP signal was acquired for the following 20 hours using a CLARIOStar Plus (BMG LABTECH) equipped with ACU to maintain 37°C and 5% CO_2_. Data were analysed using MARS software and plotted with GraphPad prims 9 software. At 22 hs post-infection, cells were fixed in 8% formaldehyde, permeabilized with 0.1 % Triton X-100 and stained with sheep anti-SARS-CoV-2 N (1:500) antiserum ^69^ followed by Alexa Fluor 594 Donkey anti-sheep IgG (H+L) (1:500, Invitrogen) and DAPI (1:4000, Sigma). Cell imagines were acquired using EVOS Cell Imaging Systems (Thermo Fischer Scientific).

### Demographic data

Data for the EVADE study were available using the NHS Greater Glasgow and Clyde (NHS GG&C) SafeHaven platform and included vaccination status (dates and product names for each dose), demographic data (age, sex and Scottish Index of Multiple Deprivation (SIMD) quartile) comorbidity (shielding and immunosuppression status) and dates of positive and negative PCR tests, for 1.2 million inhabitants of the (NHS GG&C) area over 18 years of age, from 1^st^ March 2020 up to 12^th^ January 2022. Data were matched by CHI number and pseudonymised before analysis. Derogated ethical approval was granted by the NHS GG&C SafeHaven committee (GSH/21/IM/001).

### Vaccine effectiveness

We used a logistic additive regression model to estimate relative vaccine effectiveness against the Omicron variant as it emerged in a population of 1.2 million people in NHS Greater Glasgow & Clyde, the largest health board in Scotland. Infection status for Omicron and Delta was modelled by number and product type of vaccine doses, previous infection status, sex, SIMD quartile, age on 31st October 2021 and time since most recent vaccination.

We identified Omicron infections using 3 data streams: confirmed S gene target failure (SGTF), allele specific PCR, and Pango lineage assignments from the sequencing data. SGTF samples with Delta lineage assignments were assigned as Delta infections. Samples for which the sequencing date was more than two weeks away from the first positive PCR were removed from the analysis.

We removed a small number of individuals who received ChAdOx1 as a third dose or had their third dose before the first of September 2021 on the assumption that the majority were part of the COV-BOOST clinical trial, the results of which are published elsewhere. We removed anyone with ambiguous vaccination status or whose brand was unknown due to data entry error. We removed those who were vaccinated during the study period. We removed individuals who tested positive in the 90 days before the study period. To control for the effect of missed vaccinations due to recent infection, we exclude those who were eligible for a second or third dose but had not taken this up (i.e had a first dose more than 8 weeks ago but no second dose, or a second dose more than 12 weeks ago but no third dose). Since those who tested positive during the study period could not subsequently be vaccinated within the study period, and those who changed vaccination status during the study period were excluded from the dataset, there would be an inflated proportion of people testing positive and having only a second dose of a vaccine. Specifically, those who were given the third dose of a vaccine during the study period would be excluded from the analysis, while those who would have been given a third dose of the vaccine but could not due to becoming infected would be included in the analysis. This would lead to reduced estimates of effectiveness of second doses if not accounted for appropriately. Due to the timing of the rollout of booster doses coinciding with high levels of infection, it is vital to account for this.

### Serum samples

Serum samples were collected from healthy participants in the COVID-19 Deployed Vaccine Cohort Study (DOVE), a cross-sectional post-licensing cohort study to determine the immunogenicity of deployed COVID-19 vaccines against evolving SARS-CoV-2 variants. 308 adult volunteers aged at least 18 years and were recruited into the study 14 days or more after a second or third dose of vaccine. All participants gave written informed consent to take part in the study. The DOVE study was approved by the North-West Liverpool Central Research Ethics Committee (REC reference 21/NW/0073).

### Structural modelling

The file 6vsb_1_1_1.pdb containing a complete model of the full-length glycosylated spike homotrimer in open conformation with one monomer having the receptor-binding domain in the ‘up’ position was obtained from the CHARMM-GUI Archive ^77, 78^. This model is itself generated based upon a partial spike cryo-EM structure (PDB ID: 6VSB). For visualisation, the model was trimmed to the ectodomain (residues 14-1164) and the signal peptide (residues 1-13) and glycans were removed. Using this structural model and the closed conformation equivalent (6vxx_1_1_1.pdb). Residues belonging to the receptor-binding site were identified as those with an atom within 4Å of an ACE2 atom in the bound RBD-ACE2 structure (PDB ID: 6M0J^79^) and Alpha carbon-to-Alpha carbon distances between these residues in the ‘up’ RBD and all other spike residues were calculated. Antibody accessibility scores for open and closed conformations were calculated using BEpro ^31^. Figures were prepared using PyMol ^80^.

### Epidemiological description of the emergence of the Omicron variant in the UK

On the 27^th^ November 2021, the UK Health Security Agency detected 2 cases of Omicron in England, the following day 6 Scottish cases were detected by community (Pillar 2) sequencing. Over the next 10 days (to 8^th^ December 2021) a further 95 genome sequences were obtained. Due to the rapid spread of Omicron and low genetic diversity, the genome sequences are highly related with mean genetic divergence of 1 single nucleotide polymorphisms (SNPs) and maximum 7 SNPs.

The phylogenetic relationship to Omicron sequences from other countries is consistent with multiple introductions associated with travel to South Africa followed by community transmissions within Scotland. Amongst the Scottish samples diverged from the tree backbone, there were a number identified that are genetically divergent, i.e., greater than 2 single nucleotide polymorphisms from the nearest Scottish sample (Fig. 1d). Moreover, comparison to the wider international collection of Omicron samples revealed that they were more closely related to genomes from other countries than other Scottish samples. These samples therefore likely represent independent introductions to Scotland, but without more detailed epidemiological data, the number of introductions is unknown. Where there are indistinguishable samples in the phylogeny from Scotland and elsewhere in world, importation cannot be ruled out as a source of these samples in Scotland, rather than transmission from an established population circulating in Scotland.

Within Scotland, cases are spread across 9 separate Health Boards and distributed throughout the phylogeny (Fig. 1d). Basal Scottish genomes were sampled in 7 different Health Boards, most of them from NHS Greater Glasgow & Clyde (47%) and NHS Lanarkshire (25%). Notably, amongst these earliest samples are cases that were epidemiologically linked to early spreading events. All but one of these samples were found on this basal branch and are indistinguishable, and which is consistent with transmission at these events.

### Data and materials availability

The experimental data that support the findings of this study are available on reasonable request but restrictions apply to the availability of clinical data, which were used under ethical approvals for the current study, and so are not publicly available.

Codes used in this analysis are available in the study’s GitHub repository: https://github.com/centre-for-virus-research/Omicron.

## Data Availability

All sequence data are published via GISAID.
All data produced in the present study are available upon reasonable request to the authors other than identifying clinical information.

## Acknowledgments

The authors would like to thank the participants of the DOVE study and Sister Therese McSorley and her nursing team at the NHS GG&C clinical research facility. The authors also thank Alison Hamilton, Laura Stirling and Charlie Mayor from the NHS GG&C SafeHaven team for their invaluable input in facilitating this study. We thank Paula Olmo for administrative support and Chris Robertson and Aziz Sheikh for statistical advice. The authors thank all of the researchers who have shared genome data openly via the Global Initiative on Sharing All Influenza Data (GISAID).

## Funding

Health Data Research UK (HDR UK) - the Evaluation of Variants Affecting Deployed COVID-19 Vaccine (EVADE) study (ECT, SR, OM, CW, BJW) (grant code: 2021.0155).

COG-UK is supported by funding from the Medical Research Council (MRC) part of UK Research & Innovation (UKRI), the National Institute of Health Research (NIHR) [grant code: MC_PC_19027], and Genome Research Limited, operating as the Wellcome Sanger Institute (RMB, DLR, ECT).

Medical Research Council (MRC) - the COVID-19 DeplOyed VaccinE (DOVE) study (grant code: MCUU1201412) and COG-UK (ECT).

Medical Research Council (MRC) (grant code: MC_UU_12014/12) (AF, JH, RO, JG, ECT and DLR)

Medical Research Council (MRC) (grant codes: MR/R024758/1 and MR/W005611/1) (WTH)

UK Research and Innovation (UKRI) - G2P-UK National Virology Consortium (MR/W005611/1) (MP, ECT, AP, DLR).

Wellcome Trust (grant code: 220977/Z/20/Z) (DLR)

Biotechnology and Biological Sciences Research Council (grant code: BBSRC, BB/R004250/1) (NL, BJW)

Wellcome Trust and Royal Society Sir Henry Dale Fellowship (grant code: 107653/Z/15/A) (JG)

## Author contributions

**Conceptualization:** ECT, JG, MP, DLR, BW

**Methodology:** BJW, JG, OAM, CW, NL, GL, WF, SS, MM, AS, WTH, CD, RO, JH, DP, KP, AF, GY, SS, RMP, PRM, AHP, JH, DLR, SR

**Investigation:** BJW, JG, OAM, CW, NL, GL, WF, SS, MM, AS, SA, EV, WTH, CD, RO, JH, PH, VS, DP, KP, AF, GY, SS, MTGH, RG, KT,AHP, JH,DLR,MP,SR

**Visualization:** BJW, JG, OAM, CW, GL, WTH, DLR

**Funding acquisition:** ECT, DLR, SR, BJW, JG

**Project administration:** ECT, DLR, SR, JH,

**Supervision:** BJW, DLR, MP, SR, JH, ECT

**Writing – original draft:** ECT, BJW, JG, GY

**Writing – review & editing:** BJW, JG, OAM, CW, SA, WTH, DLR, MP, ECT

## Data and materials availability

The experimental data that support the findings of this study are available on reasonable request but restrictions apply to the availability of clinical data, which were used under ethical approvals for the current study, and so are not publicly available. Anonymised data are available with permission of the NHS Greater Glasgow & Clyde SafeHaven.

Codes used in this analysis are available upon request from the corresponding author.

Biological materials including cell lines are available on reasonable request from the authors. Clinical samples are restricted for use under the ethical approvals obtained for their use.

## Acknowledgments

The authors would like to thank the participants of the DOVE study and Sister Therese McSorley and her nursing team at the NHS GG&C clinical research facility. The authors also thank Alison Hamilton, Laura Stirling and Charlie Mayor from the NHS GG&C SafeHaven team for their invaluable input in facilitating this study. We thank Paula Olmo for administrative support and Chris Robertson and Aziz Sheikh for statistical advice. The authors thank all of the researchers who have shared genome data openly via the Global Initiative on Sharing All Influenza Data (GISAID). Funding was provided by Health Data Research UK (HDR UK) for the Evaluation of Variants Affecting Deployed COVID-19 Vaccine (EVADE) study (ECT, SR, OM, CW, BJW, grant code: 2021.0155). This research is part of the Data and Connectivity National Core Study, led by Health Data Research UK in partnership with the Office for National Statistics and funded by UK Research and Innovation (grant ref MC_PC_20058). This work was also supported by The Alan Turing Institute via ‘Towards Turing 2.0’ EPSRC Grant Funding. COG-UK is supported by funding from the Medical Research Council (MRC, part of UK Research & Innovation (UKRI)), the National Institute of Health Research (NIHR) [grant code: MC_PC_19027], and Genome Research Limited, operating as the Wellcome Sanger Institute (RMB, DLR, ECT). Medical Research Council (MRC) provided funding for both the COVID-19 DeplOyed VaccinE (DOVE) study (grant code: MCUU1201412) and COG-UK (ECT). AF, JH, RO, JG, ECT, NL and DLR were funded by the Medical Research Council (MRC) (grant code: MC_UU_12014/12). W.T.H. was supported by Medical Research Council (MRC), grant codes MR/R024758/1 and MR/W005611/1. The G2P-UK National Virology Consortium was funded by UK Research and Innovation (UKRI) award MR/W005611/1 (MP, ECT, AP, DLR). D.L.R. was funded by Wellcome Trust (grant code: 220977/Z/20/Z). N.L. and B.J.W. were funded by the Biotechnology and Biological Sciences Research Council (grant codes: BBSRC, BB/R004250/1 & BB/R019843/1). J.G. was funded by a Wellcome Trust and Royal Society Sir Henry Dale Fellowship (grant code: 107653/Z/15/A).

## Author information

These authors contributed equally: Brian J. Willett, Joe Grove, Oscar A. MacLean, Craig Wilkie, Nicola Logan, Giuditta De Lorenzo & Wilhelm Furnon

These authors contributed equally: John Haughney, David L. Robertson, Massimo Palmarini, Surajit Ray & Emma C. Thomson

## Contributions

**Conceptualization:** ECT, JG, MP, DLR, BW

**Investigation:** BJW, JG, OAM, CW, NL, GL, WF, SS, MM, AS, SA, EV, WTH, CD, RO, JH, PH, VS, DP, KP, AF, GY, SS, MTGH, RG, KT, AHP, JH, DLR, MP, SR

**Visualization:** BJW, JG, OAM, CW, GL, WTH, DLR

**Funding acquisition:** ECT, DLR, SR, BJW, JG

**Project administration:** ECT, DLR, SR, JH

**Supervision:** BJW, DLR, MP, SR, JH, ECT

**Writing – original draft:** ECT, BJW, JG, GY

**Writing – review & editing:** BJW, JG, OAM, CW, SA, WTH, DLR, MP, ECT

## Corresponding authors

Correspondence to Professor Emma Thomson and Professor Brian Willett, MRC-University of Glasgow Centre for Virus Research, UK, G61 1QH.

## Ethics declarations

The authors declare that they have no competing interests.

## Extended Data

**Extended data Fig.1.**
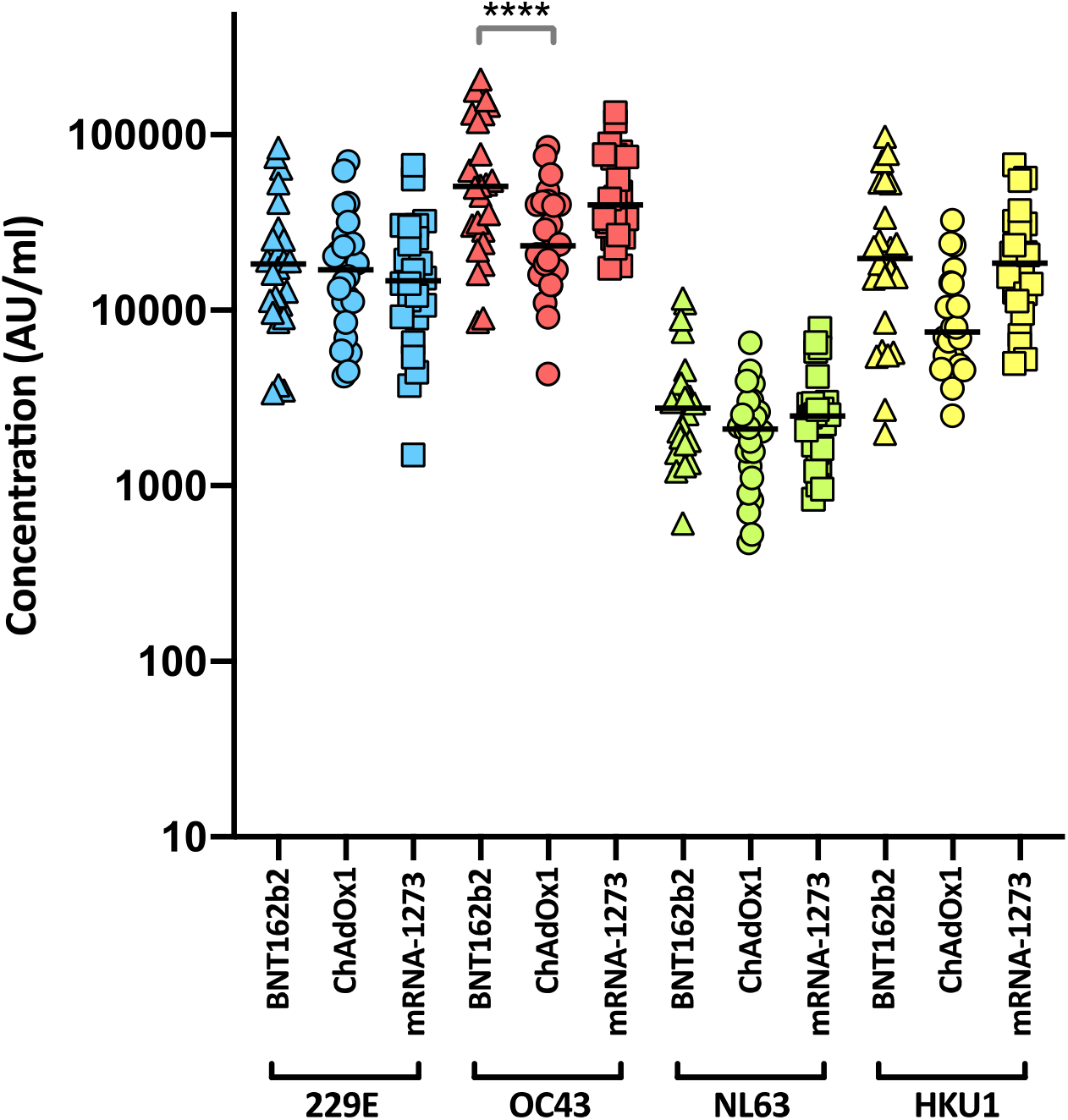
HCoV reactivity following two doses of SARS-CoV-2 vaccine. Antibody responses were studied in three groups of individuals (n=24 per group) vaccinated with either BNT162b2, ChAdOx1 or mRNA-1273 by MSD-ECL assay. Responses were measured against full-length spike glycoprotein (Spike) from HCoVs 229E, OC43, NL63 and HKU1 and are expressed as MSD arbitrary units (AU/ml). The response to OC43 was significantly higher in BNT162b2 vaccinates than in ChAdOx1 vaccinates.

**Extended data Fig. 2.**
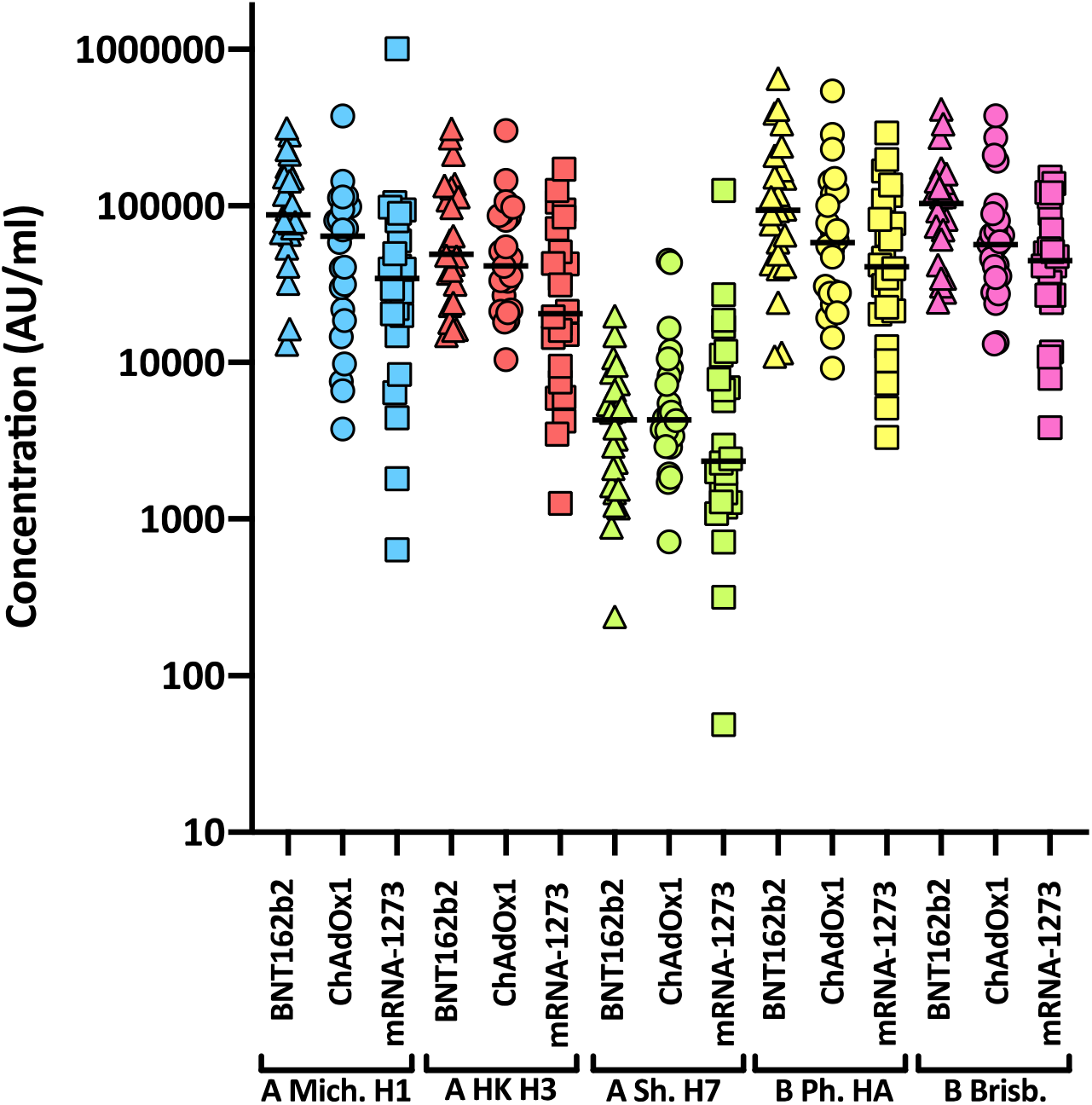
Influenza reactivity following two doses of SARS-CoV-2 vaccine. Antibody responses were studied in three groups of individuals (n=24 per group) vaccinated with either BNT162b2, ChAdOx1 or mRNA-1273 by MSD-ECL assay. Responses were measured against haemagglutinins from influenza viruses; influenza A Michigan H1, Hong Kong H3 and Shanghai H7, and influenza B Phuket HA and Brisbane and are expressed as MSD arbitrary units (AU/ml). No significant differences were detected between the vaccine groups for each of the antigens.

**Extended data Fig. 3.**
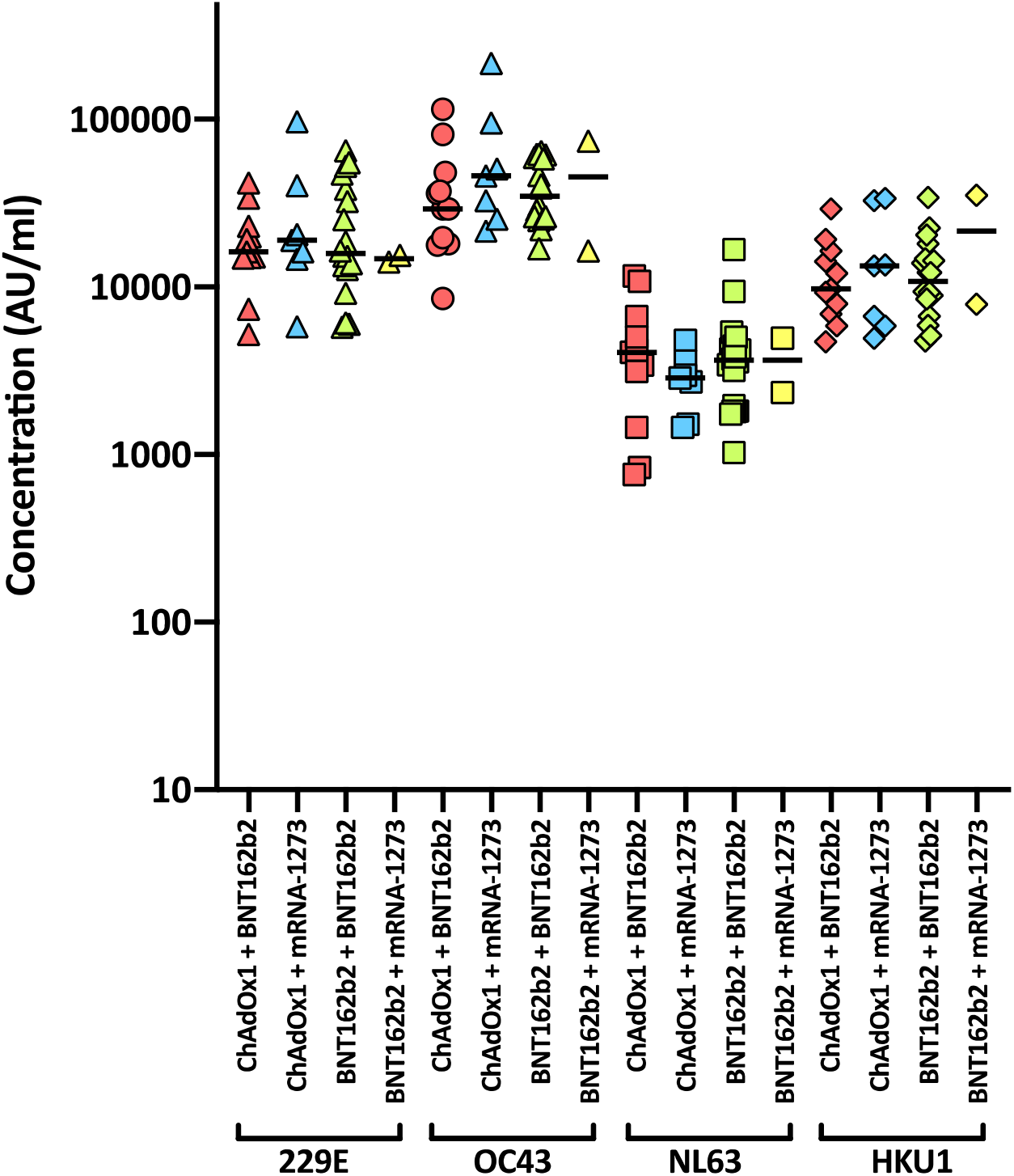
HCoV reactivity following third dose of SARS-CoV-2 vaccine. Antibody responses were studied in four groups of individuals primed with two doses of either ChAdOx1 or BNT162b2, followed by a booster of BNT162b2 or mRNA-1273. Responses were measured by MSD-ECL assay against full-length spike glycoprotein (Spike) from HCoVs 229E, OC43, NL63 and HKU1 and are expressed as MSD arbitrary units (AU/ml).

**Extended data Fig. 4.**
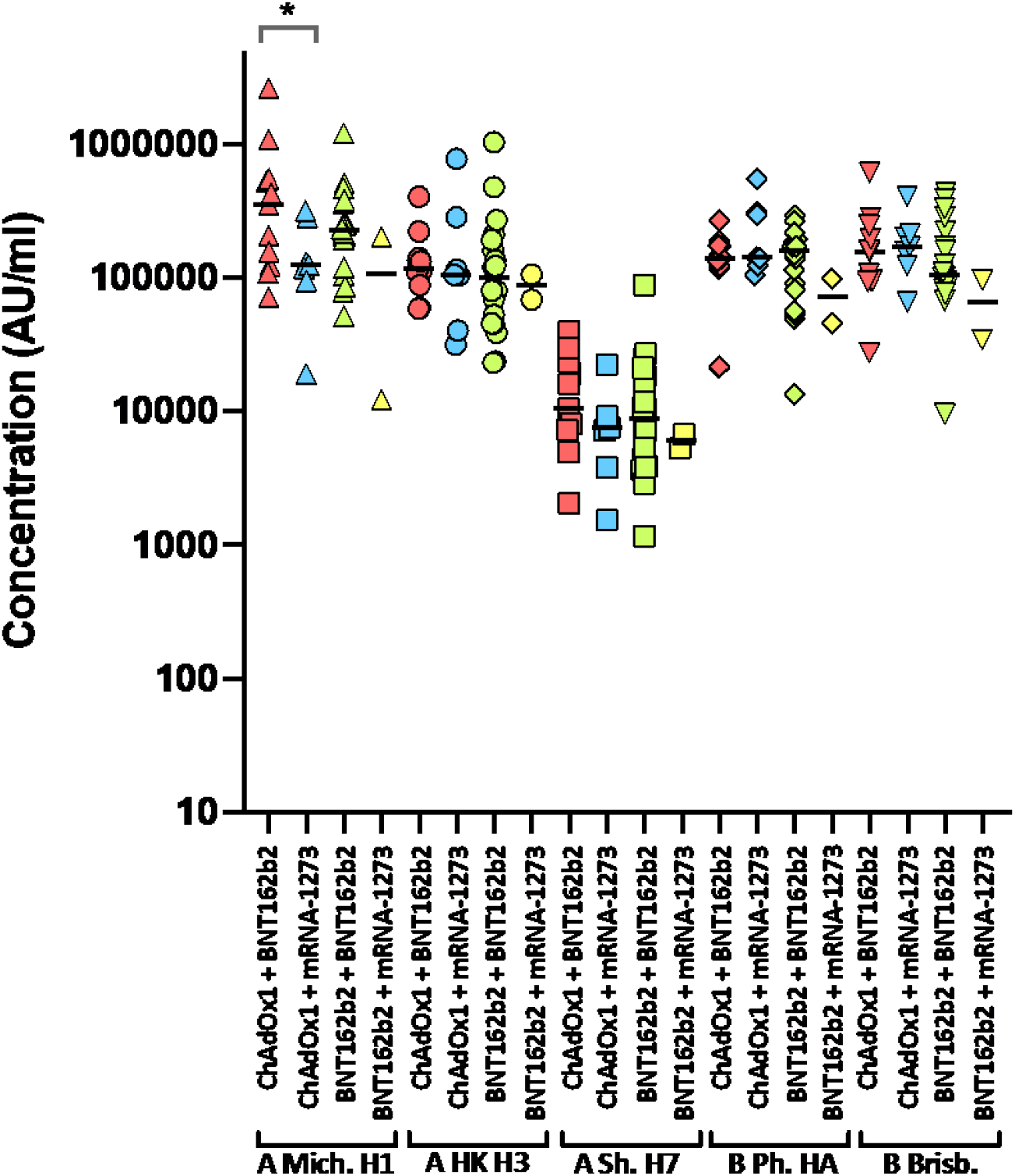
Influenza reactivity following third dose of SARS-CoV-2 vaccine. Antibody responses were studied in four groups of individuals primed with two doses of either ChAdOx1 or BNT162b2, followed by a booster of BNT162b2 or mRNA-1273. Responses were measured by MSD-ECL against haemagglutinins from influenza viruses; influenza A Michigan H1, Hong Kong H3 and Shanghai H7, and influenza B Phuket HA and Brisbane and are expressed as MSD arbitrary units (AU/ml). * Significantly different p=0.0413.

**Extended Data Fig. 5.**
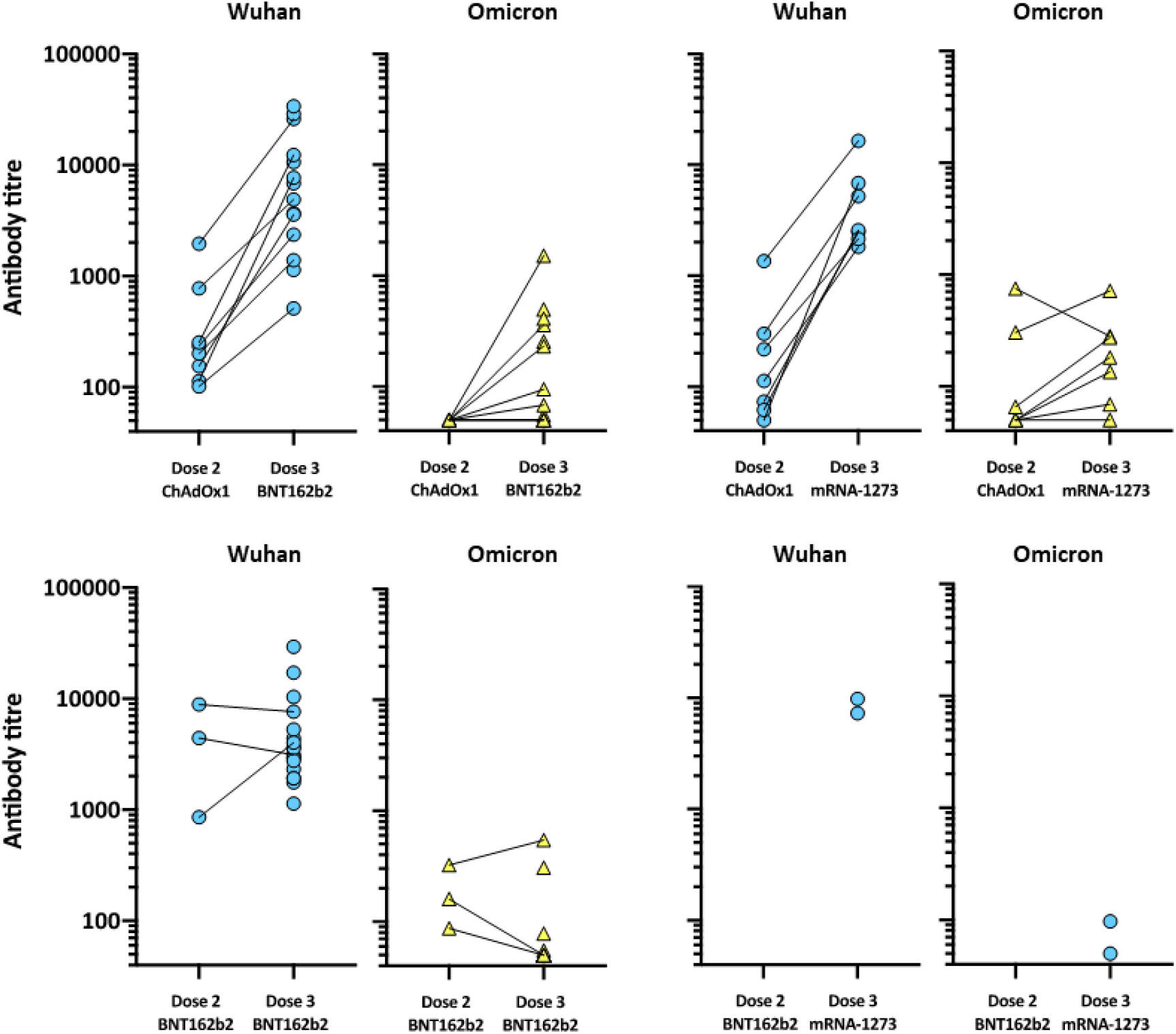
Effect of third dose of SARS-CoV-2 vaccine on neutralising antibody titres. Two groups of healthy volunteers vaccinated with two doses of either ChAdOx1 or BNT162b2, were sampled two weeks following a third dose of either BNT162b2 or mRNA-1273. Each point represents the mean of three replicates. Where dose 2 and dose 3 samples were available from the same individual, points are joined by a solid line.

**Extended Data Fig.6.**
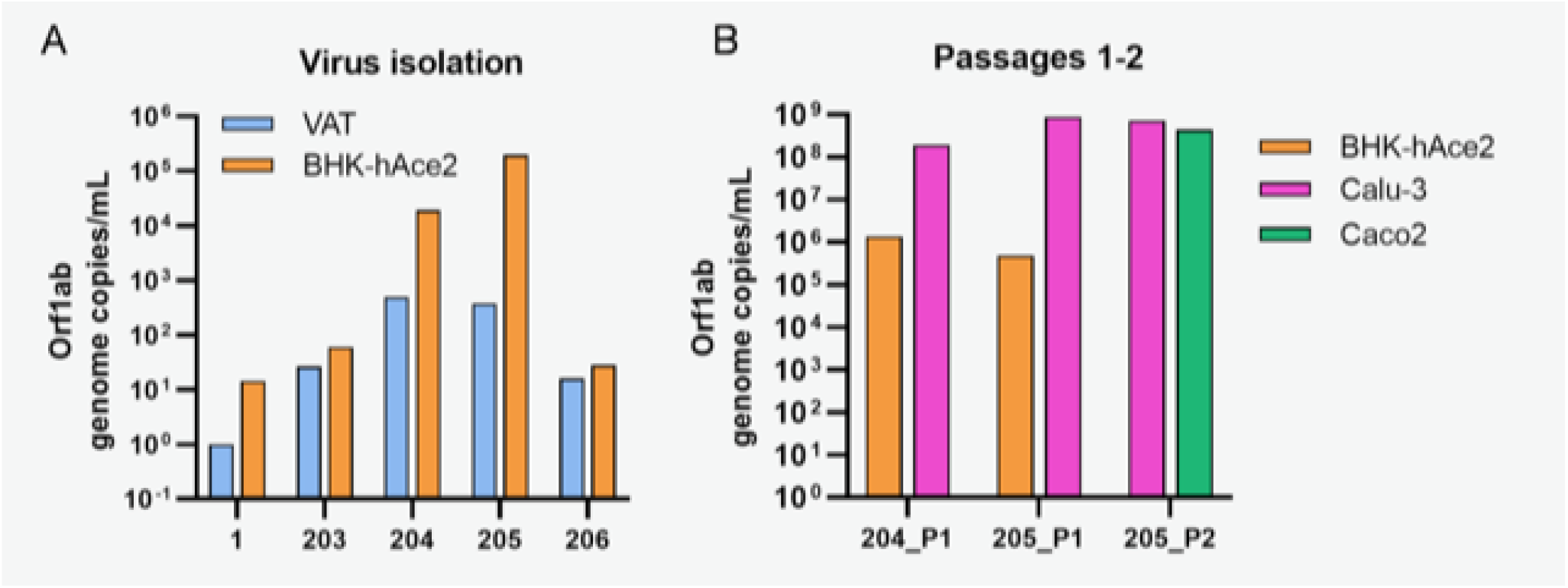
Isolation of Omicron in cell culture. **a**, Vero ACE2 TMPRSS2 (VAT) and BHK-hACE2 cells were inoculated with diluted clinical samples. Viral progeny was quantified in the medium 5 dpi by RT-qPCR. **b,** Aliquots of the medium from samples named 204 and 205 were used to generate a P1 in BHK-hACE2 and Calu-3 cells and, limited to sample 205, a P2 in Calu-3 and Caco2 cells. Viral stocks were quantified by RT-qPCR.

**Supplementary Table 1.** Comparison of SARS-CoV-2 antibody responses elicited by two doses of SARS-CoV-2 vaccine. Data were analyzed in GraphPad Prism v8.4.3, groups were compared by ordinary one-way ANOVA.

| Tukey's multiple comparisons test | Mean Diff. | 95.00% CI of diff. | Significant? | Summary | Adjusted P Value |
| --- | --- | --- | --- | --- | --- |
| Spike BNT162b2 vs. ChAdOx1 | 2588 | -278.1 to 5453 | No | ns | 0.1212 |
| Spike BNT162b2 vs. mRNA-1273 | -248.4 | -3114 to 2617 | No | ns | >0.9999 |
| Spike ChAdOx1 vs. mRNA-1273 | -2836 | -5702 to 29.73 | No | ns | 0.0553 |
| RBD BNT162b2 vs. ChAdOx1 | 4649 | 1783 to 7514 | Yes | **** | <0.0001 |
| RBD BNT162b2 vs. mRNA-1273 | -449.1 | -3315 to 2417 | No | ns | >0.9999 |
| RBD ChAdOx1 vs. mRNA-1273 | -5098 | -7964 to -2232 | Yes | **** | <0.0001 |
| NTD BNT162b2 vs. ChAdOx1 | 5755 | 2889 to 8620 | Yes | **** | <0.0001 |
| NTD BNT162b2 vs. mRNA-1273 | 714.7 | -2151 to 3581 | No | ns | 0.9996 |
| NTD ChAdOx1 vs. mRNA-1273 | -5040 | -7906 to -2174 | Yes | **** | <0.0001 |
| N BNT162b2 vs. ChAdOx1 | 2.183 | -2864 to 2868 | No | ns | >0.9999 |
| N BNT162b2 vs. mRNA-1273 | 0.4 | -2865 to 2866 | No | ns | >0.9999 |
| N ChAdOx1 vs. mRNA-1273 | -1.783 | -2868 to 2864 | No | ns | >0.9999 |

**Supplementary Table 2.** Comparison of HCoV antibody responses elicited by two doses of SARS-CoV-2 vaccine. Data were analyzed in GraphPad Prism v8.4.3, groups were compared by ordinary one-way ANOVA.

| Tukey's multiple comparisons test | Mean Diff. | 95.00% CI of diff. | Significant? | Summary | Adjusted P Value |
| --- | --- | --- | --- | --- | --- |
| 229E BNT162b2 vs. ChAdOx1 | 3296 | -19673 to 26265 | No | ns | >0.9999 |
| 229E BNT162b2 vs. mRNA-1273 | 5282 | -17687 to 28251 | No | ns | 0.9998 |
| 229E ChAdOx1 vs. mRNA-1273 | 1986 | -20982 to 24955 | No | ns | >0.9999 |
| OC43 BNT162b2 vs. ChAdOx1 | 39581 | 16612 to 62550 | **Yes** | **\*\*\*\*** | **<0.0001** |
| OC43 BNT162b2 vs. mRNA-1273 | 18396 | -4573 to 41365 | No | ns | 0.2628 |
| OC43 ChAdOx1 vs. mRNA-1273 | -21185 | -44154 to 1784 | No | ns | 0.1027 |
| NL63 BNT162b2 vs. ChAdOx1 | 1317 | -21652 to 24286 | No | ns | >0.9999 |
| NL63 BNT162b2 vs. mRNA-1273 | 696.9 | -22272 to 23666 | No | ns | >0.9999 |
| NL63 ChAdOx1 vs. mRNA-1273 | -620 | -23589 to 22349 | No | ns | >0.9999 |
| HKU1 BNT162b2 vs. ChAdOx1 | 17914 | -5055 to 40883 | No | ns | 0.3015 |
| HKU1 BNT162b2 vs. mRNA-1273 | 5468 | -17501 to 28437 | No | ns | 0.9998 |
| HKU1 ChAdOx1 vs. mRNA-1273 | -12446 | -35415 to 10523 | No | ns | 0.8243 |

**Supplementary Table 3.**
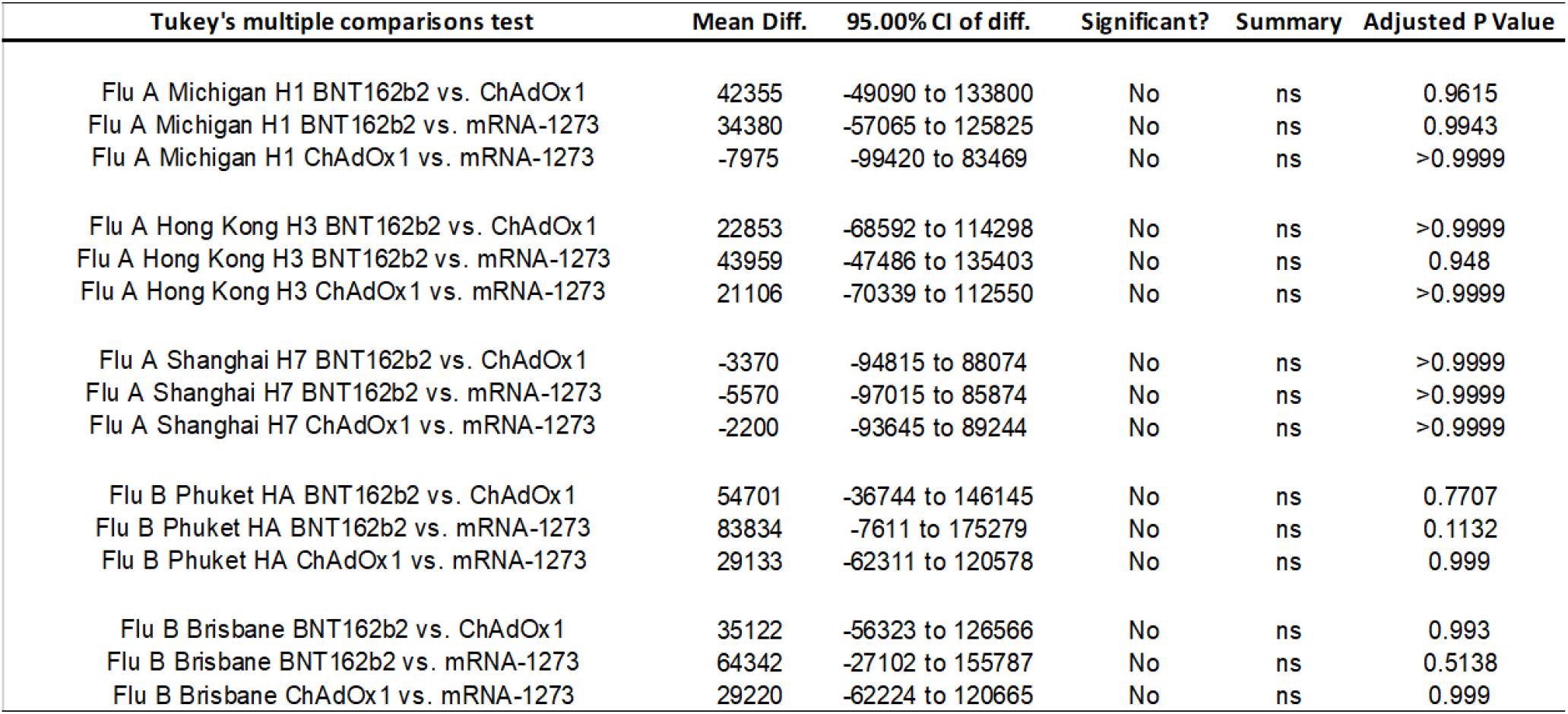
Comparison of influenza antibody responses elicited by two doses of SARS-CoV-2 vaccine. Data were analyzed in GraphPad Prism v8.4.3, groups were compared by ordinary one-way ANOVA.

**Supplementary Table 4.** Comparison of neutralising antibody titres elicited by two doses of SARS-CoV-2 vaccine. Neutralising antibody responses were quantified against Wuhan or Omicron spike glycoprotein-bearing HIV(SARS-CoV-2) pseudotypes. Data were analyzed in GraphPad Prism v8.4.3, groups were compared by ordinary one-way ANOVA.

| Tukey's multiple comparisons test | Mean 1 | Mean 2 | Mean Diff. | 95.00% CI of diff. | Significant? | Summary | Adjusted P Value |
| --- | --- | --- | --- | --- | --- | --- | --- |
| Wuhan BNT162b2 vs. ChAdOx1 | 4978 | 882.3 | 4096 | 733.8 to 7458 | Yes | ** | 0.0075 |
| Wuhan BNT162b2 vs. mRNA-1273 | 4978 | 21118 | -16140 | -19502 to -12778 | Yes | **** | <0.0001 |
| Wuhan ChAdOx1 vs. mRNA-1273 | 882.3 | 21118 | -20236 | -23598 to -16874 | Yes | **** | <0.0001 |
| Omicron BNT162b2 vs. ChAdOx1 | 148.3 | 61.9 | 86.39 | -3276 to 3448 | No | ns | >0.9999 |
| Omicron BNT162b2 vs. mRNA-1273 | 148.3 | 285 | -136.7 | -3499 to 3225 | No | ns | >0.9999 |
| Omicron ChAdOx1 vs. mRNA-1273 | 61.9 | 285 | -223.1 | -3585 to 3139 | No | ns | >0.9999 |
| BNT162b2 Wuhan vs. Omicron | 4978 | 148.3 | 4830 | 1468 to 8192 | Yes | *** | 0.0008 |
| ChAdOx12 Wuhan vs. Omicron | 882.3 | 61.9 | 820.4 | -2542 to 4182 | No | ns | 0.981 |
| mRNA-1273 Wuhan vs. Omicron | 21118 | 285 | 20833 | 17471 to 24195 | Yes | **** | <0.0001 |

**Supplementary Table 5.** Comparison of SARS-CoV-2 antibody responses elicited by a third dose of SARS-CoV-2 vaccine. Data were analyzed in GraphPad Prism v8.4.3, groups were compared by ordinary one-way ANOVA. P= BNT162b2, AZ = ChAdOx1, M = mRNA-1273.

| Tukey's multiple comparisons test | Mean Diff. | 95.00% CI of diff. | Significant? | Summary | Adjusted P Value |
| --- | --- | --- | --- | --- | --- |
| Spike AZ+P vs. AZ+M | 1163 | -4354 to 6680 | No | ns | >0.9999 |
| Spike AZ+P vs. P+P | 1007 | -3360 to 5374 | No | ns | >0.9999 |
| Spike AZ+P vs. P+M | 361.3 | -8410 to 9133 | No | ns | >0.9999 |
| Spike AZ+M vs. P+P | -156 | -5239 to 4927 | No | ns | >0.9999 |
| Spike AZ+M vs. P+M | -801.5 | -9951 to 8348 | No | ns | >0.9999 |
| Spike P+P vs. P+M | -645.5 | -9151 to 7860 | No | ns | >0.9999 |
| RBD AZ+P vs. AZ+M | 2675 | -2842 to 8192 | No | ns | 0.9453 |
| RBD AZ+P vs. P+P | 1439 | -2928 to 5806 | No | ns | 0.9988 |
| RBD AZ+P vs. P+M | 37.5 | -8734 to 8809 | No | ns | >0.9999 |
| RBD AZ+M vs. P+P | -1236 | -6319 to 3846 | No | ns | >0.9999 |
| RBD AZ+M vs. P+M | -2638 | -11787 to 6511 | No | ns | 0.9997 |
| RBD P+P vs. P+M | -1401 | -9906 to 7104 | No | ns | >0.9999 |
| NTD AZ+P vs. AZ+M | 3368 | -2149 to 8885 | No | ns | 0.7418 |
| NTD AZ+P vs. P+P | 4098 | -269.4 to 8465 | No | ns | 0.0919 |
| NTD AZ+P vs. P+M | 3739 | -5033 to 12510 | No | ns | 0.9822 |
| NTD AZ+M vs. P+P | 729.8 | -4353 to 5813 | No | ns | >0.9999 |
| NTD AZ+M vs. P+M | 370.8 | -8778 to 9520 | No | ns | >0.9999 |
| NTD P+P vs. P+M | -359 | -8864 to 8146 | No | ns | >0.9999 |
| N AZ+P vs. AZ+M | 2.755 | -5514 to 5520 | No | ns | >0.9999 |
| N AZ+P vs. P+P | -0.7066 | -4368 to 4366 | No | ns | >0.9999 |
| N AZ+P vs. P+M | -3.095 | -8775 to 8768 | No | ns | >0.9999 |
| N AZ+M vs. P+P | -3.461 | -5086 to 5079 | No | ns | >0.9999 |
| N AZ+M vs. P+M | -5.85 | -9155 to 9143 | No | ns | >0.9999 |
| N P+P vs. P+M | -2.389 | -8508 to 8503 | No | ns | >0.9999 |

P = BNT162b2; AZ = ChAdOx1; M = mRNA-1273

**Supplementary Table 6.** Comparison of HCoV antibody responses elicited by a third dose of SARS-CoV-2 vaccine. Data were analyzed in GraphPad Prism v8.4.3, groups were compared by ordinary one-way ANOVA. P= BNT162b2, AZ = ChAdOx1, M = mRNA-1273.

| Tukey's multiple comparisons test | Mean Diff. | 95.00% CI of diff. | Significant? | Summary | Adjusted P Value |
| --- | --- | --- | --- | --- | --- |
| 229E AZ+P vs. AZ+M | -11229 | -47020 to 24562 | No | ns | 0.9993 |
| 229E AZ+P vs. P+P | -5670 | -34000 to 22661 | No | ns | >0.9999 |
| 229E AZ+P vs. P+M | 4487 | -52417 to 61391 | No | ns | >0.9999 |
| 229E AZ+M vs. P+P | 5560 | -27414 to 38533 | No | ns | >0.9999 |
| 229E AZ+M vs. P+M | 15716 | -43637 to 75069 | No | ns | >0.9999 |
| 229E P+P vs. P+M | 10156 | -45019 to 65332 | No | ns | >0.9999 |
| OC43 AZ+P vs. AZ+M | -29568 | -65359 to 6224 | No | ns | 0.2369 |
| OC43 AZ+P vs. P+P | -121 | -28451 to 28209 | No | ns | >0.9999 |
| OC43 AZ+P vs. P+M | -5301 | -62205 to 51603 | No | ns | >0.9999 |
| OC43 AZ+M vs. P+P | 29447 | -3527 to 62420 | No | ns | 0.1382 |
| OC43 AZ+M vs. P+M | 24266 | -35086 to 83619 | No | ns | 0.988 |
| OC43 P+P vs. P+M | -5180 | -60356 to 49996 | No | ns | >0.9999 |
| NL63 AZ+P vs. AZ+M | -11140 | -46931 to 24651 | No | ns | 0.9994 |
| NL63 AZ+P vs. P+P | -8074 | -36404 to 20256 | No | ns | 0.9998 |
| NL63 AZ+P vs. P+M | -16842 | -73746 to 40062 | No | ns | 0.9996 |
| NL63 AZ+M vs. P+P | 3066 | -29908 to 36040 | No | ns | >0.9999 |
| NL63 AZ+M vs. P+M | -5702 | -65055 to 53651 | No | ns | >0.9999 |
| NL63 P+P vs. P+M | -8768 | -63944 to 46407 | No | ns | >0.9999 |
| HKU1 AZ+P vs. AZ+M | 9482 | -26309 to 45273 | No | ns | >0.9999 |
| HKU1 AZ+P vs. P+P | 8028 | -20303 to 36358 | No | ns | 0.9998 |
| HKU1 AZ+P vs. P+M | 8678 | -48226 to 65582 | No | ns | >0.9999 |
| HKU1 AZ+M vs. P+P | -1454 | -34428 to 31519 | No | ns | >0.9999 |
| HKU1 AZ+M vs. P+M | -804.1 | -60157 to 58549 | No | ns | >0.9999 |
| HKU1 P+P vs. P+M | 650.4 | -54525 to 55826 | No | ns | >0.9999 |

P = BNT162b2; AZ = ChAdOx1; M = mRNA-1273

**Supplementary Table 7.** Comparison of influenza antibody responses elicited by a third dose of SARS-CoV-2 vaccine. Data were analyzed in GraphPad Prism v8.4.3, groups were compared by ordinary one-way ANOVA. P= BNT162b2, AZ = ChAdOx1, M = mRNA-1273.

| Tukey's multiple comparisons test | Mean Diff. | 95.00% CI of diff. | Significant? | Summary | Adjusted P Value |
| --- | --- | --- | --- | --- | --- |
| Flu A Michigan H1 AZ+P vs. AZ+M | 410671 | 6882 to 814460 | Yes | * | 0.0413 |
| Flu A Michigan H1 AZ+P vs. P+P | 279008 | -40609 to 598625 | No | ns | 0.1757 |
| Flu A Michigan H1 AZ+P vs. P+M | 459436 | -182547 to 1101420 | No | ns | 0.5271 |
| Flu A Michigan H1 AZ+M vs. P+P | -131663 | -503667 to 240341 | No | ns | 0.9993 |
| Flu A Michigan H1 AZ+M vs. P+M | 48766 | -620842 to 718373 | No | ns | >0.9999 |
| Flu A Michigan H1 P+P vs. P+M | 180429 | -442054 to 802911 | No | ns | >0.9999 |
| Flu A Hong Kong H3 AZ+P vs. AZ+M | -64025 | -467814 to 339763 | No | ns | >0.9999 |
| Flu A Hong Kong H3 AZ+P vs. P+P | -32804 | -352421 to 286812 | No | ns | >0.9999 |
| Flu A Hong Kong H3 AZ+P vs. P+M | 56571 | -585413 to 698554 | No | ns | >0.9999 |
| Flu A Hong Kong H3 AZ+M vs. P+P | 31221 | -340783 to 403225 | No | ns | >0.9999 |
| Flu A Hong Kong H3 AZ+M vs. P+M | 120596 | -549012 to 790204 | No | ns | >0.9999 |
| Flu A Hong Kong H3 P+P vs. P+M | 89375 | -533107 to 711857 | No | ns | >0.9999 |
| Flu A Shanghai H7 AZ+P vs. AZ+M | 6841 | -396948 to 410630 | No | ns | >0.9999 |
| Flu A Shanghai H7 AZ+P vs. P+P | 469.8 | -319147 to 320087 | No | ns | >0.9999 |
| Flu A Shanghai H7 AZ+P vs. P+M | 9363 | -632620 to 651346 | No | ns | >0.9999 |
| Flu A Shanghai H7 AZ+M vs. P+P | -6371 | -378375 to 365633 | No | ns | >0.9999 |
| Flu A Shanghai H7 AZ+M vs. P+M | 2522 | -667086 to 672130 | No | ns | >0.9999 |
| Flu A Shanghai H7 P+P vs. P+M | 8893 | -613589 to 631376 | No | ns | >0.9999 |
| Flu B Phuket HA AZ+P vs. AZ+M | -101482 | -505271 to 302307 | No | ns | >0.9999 |
| Flu B Phuket HA AZ+P vs. P+P | -6309 | -325926 to 313308 | No | ns | >0.9999 |
| Flu B Phuket HA AZ+P vs. P+M | 65660 | -576323 to 707643 | No | ns | >0.9999 |
| Flu B Phuket HA AZ+M vs. P+P | 95173 | -276831 to 467178 | No | ns | >0.9999 |
| Flu B Phuket HA AZ+M vs. P+M | 167142 | -502466 to 836750 | No | ns | >0.9999 |
| Flu B Phuket HA P+P vs. P+M | 71969 | -550513 to 694451 | No | ns | >0.9999 |
| Flu B Brisbane AZ+P vs. AZ+M | -2954 | -406743 to 400834 | No | ns | >0.9999 |
| Flu B Brisbane AZ+P vs. P+P | 24661 | -294956 to 344278 | No | ns | >0.9999 |
| Flu B Brisbane AZ+P vs. P+M | 122461 | -519522 to 764444 | No | ns | >0.9999 |
| Flu B Brisbane AZ+M vs. P+P | 27615 | -344389 to 399620 | No | ns | >0.9999 |
| Flu B Brisbane AZ+M vs. P+M | 125415 | -544193 to 795023 | No | ns | >0.9999 |
| Flu B Brisbane P+P vs. P+M | 97800 | -524683 to 720282 | No | ns | >0.9999 |

P = BNT162b2; AZ = ChAdOx1; M = mRNA-1273

**Supplementary Table 8.** Effect of third dose of SARS-CoV-2 vaccine on neutralising antibody titres. Neutralising antibody responses were quantified against Wuhan or Omicron spike glycoprotein-bearing HIV (SARS-CoV-2) pseudotypes. Data were analyzed in GraphPad Prism v8.4.3, groups were compared by ordinary one-way ANOVA.

| BNT162b2 prime (doses 1 & 2) |  |  |  |  |  |
| --- | --- | --- | --- | --- | --- |
| Tukey's multiple comparisons test | Mean Diff. | 95.00% CI of diff. | Significant? | Summary | Adjusted P Value |
| Wuhan (dose 2) vs. (dose 3) | -1186 | -4381 to 2010 | No | ns | 0.7655 |
| Omicron (dose 2) vs. (dose 3) | 56.56 | -3139 to 3252 | No | ns | >0.9999 |
| Test details | Mean 1 | Mean 2 | Mean Diff. | SE of diff. | n1 |
| Wuhan (dose 2) vs. (dose 3) | 4978 | 6164 | -1186 | 1219 | 24 |
| Omicron (dose 2) vs. (dose 3) | 148.3 | 91.73 | 56.56 | 1219 | 24 |
| ChAdOx1 prime (doses 1 & 2) |  |  |  |  |  |
| Tukey's multiple comparisons test | Mean Diff. | 95.00% CI of diff. | Significant? | Summary | Adjusted P Value |
| Wuhan (dose 2) vs. (dose 3) | -7360 | -11073 to -3647 | Yes | **** | <0.0001 |
| Omicron (dose 2) vs. (dose 3) | -203.7 | -3916 to 3509 | No | ns | 0.9989 |
| Test details | Mean 1 | Mean 2 | Mean Diff. | SE of diff. | n1 |
| Wuhan (dose 2) vs. (dose 3) | 882.3 | 8242 | -7360 | 1416 | 24 |
| Omicron (dose 2) vs. (dose 3) | 61.9 | 265.5 | -203.7 | 1416 | 24 |
| Dose 3 titre comparison |  |  |  |  |  |
| Tukey's multiple comparisons test | Mean Diff. | 95.00% CI of diff. | Significant? | Summary | Adjusted P Value |
| Wuhan (BNT162b2 dose 2) vs. (ChAdOX1 dose 2) | -2079 | -7008 to 2851 | No | ns | 0.6859 |
| Omicron (BNT162b2 dose 2) vs. (ChAdOX1 dose 2) | -173.8 | -5103 to 4756 | No | ns | 0.9997 |
| Test details | Mean 1 | Mean 2 | Mean Diff. | SE of diff. | n1 |
| Wuhan (BNT162b2 dose 2) vs. (ChAdOX1 dose 2) | 6164 | 8242 | -2079 | 1877 | 20 |
| Omicron (BNT162b2 dose 2) vs. (ChAdOX1 dose 2) | 91.73 | 265.5 | -173.8 | 1877 | 20 |

**Supplementary Table 9:**
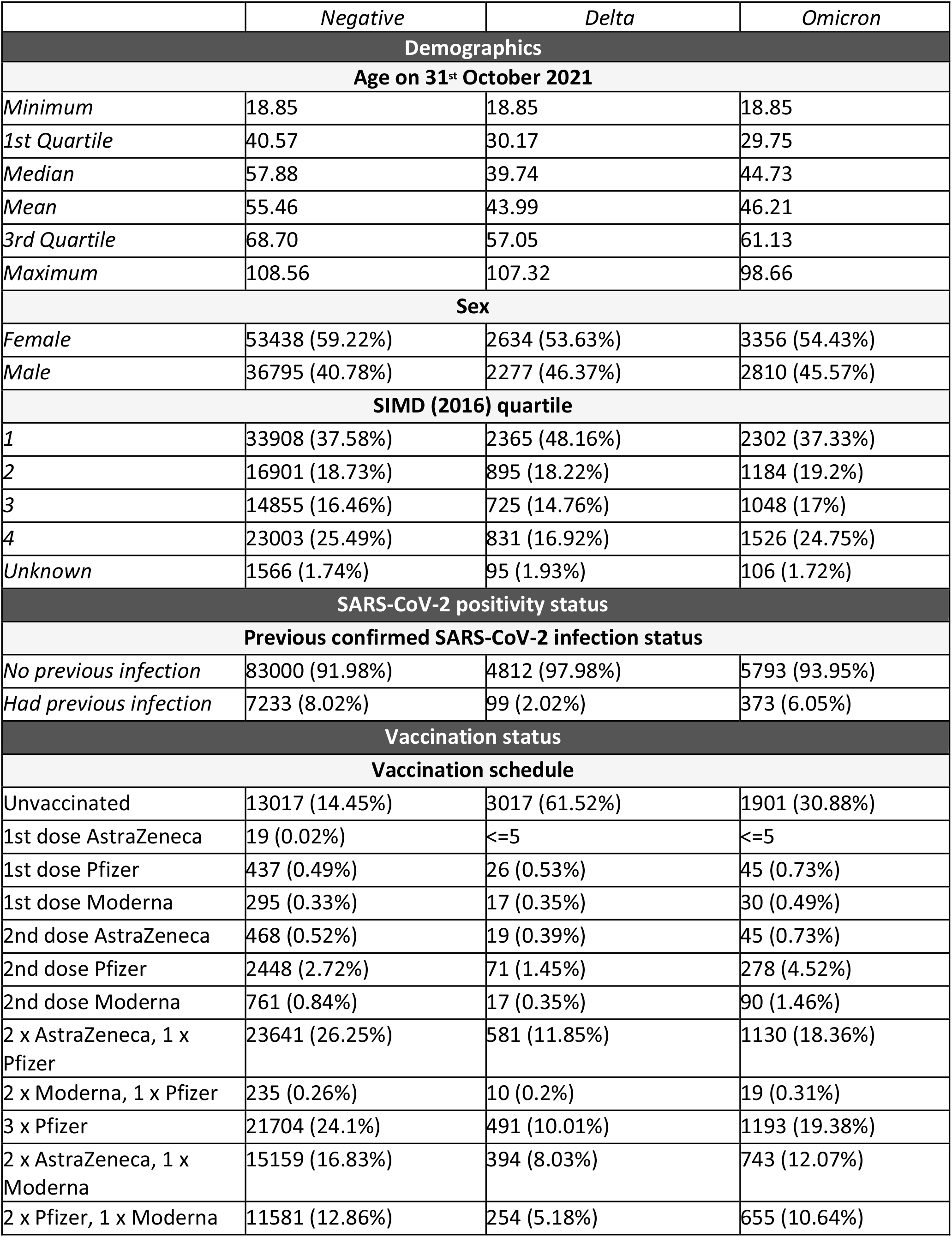
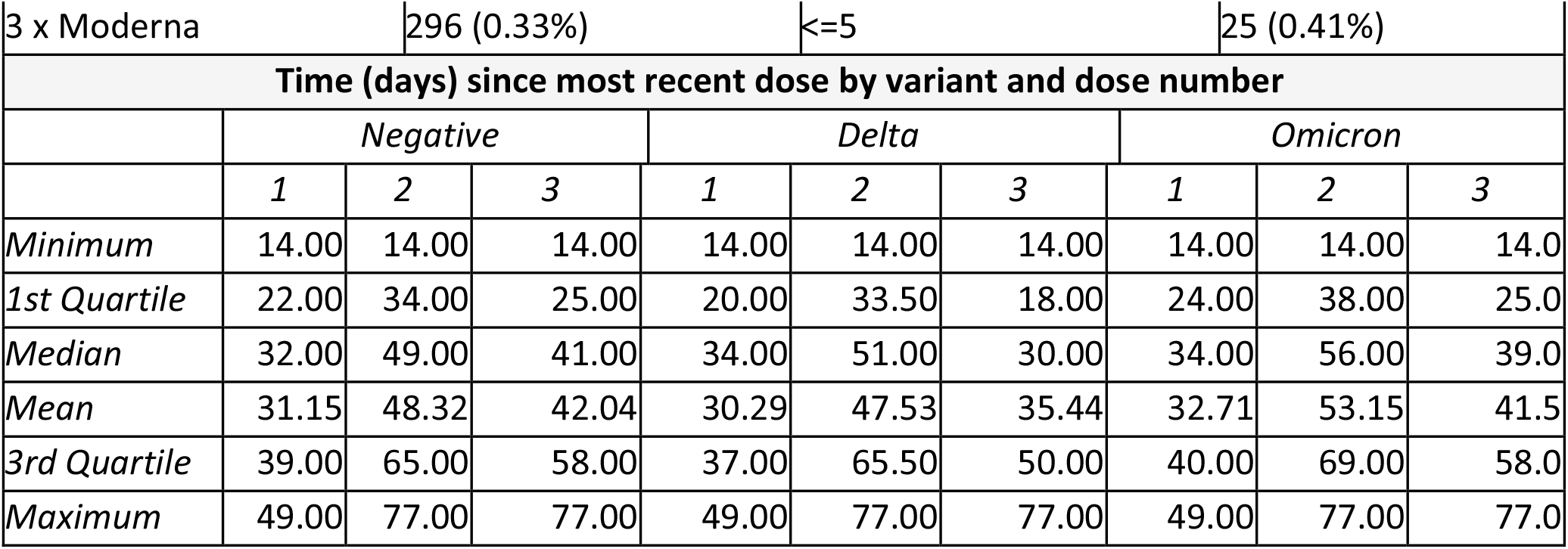
Demographics, SARS-CoV-2 positivity status and vaccination status for the study population. Population consisted of 101,310 people aged 18 and over, registered as living in NHS Greater Glasgow and Clyde and tested by PCR test for SARS-CoV-2 infection between 22nd and 28th December 2021, split by SARS-CoV-2 variant status.

|  | *Negative* | *Delta* | *Omicron* |
| --- | --- | --- | --- |
| **Demographics** |  |  |  |
| **Age on 31<sup>st</sup> October 2021** |  |  |  |
| *Minimum* | 18.85 | 18.85 | 18.85 |
| *1st Quartile* | 40.57 | 30.17 | 29.75 |
| *Median* | 57.88 | 39.74 | 44.73 |
| *Mean* | 55.46 | 43.99 | 46.21 |
| *3rd Quartile* | 68.70 | 57.05 | 61.13 |
| *Maximum* | 108.56 | 107.32 | 98.66 |
| **Sex** |  |  |  |
| *Female* | 53438 (59.22%) | 2634 (53.63%) | 3356 (54.43%) |
| *Male* | 36795 (40.78%) | 2277 (46.37%) | 2810 (45.57%) |
| **SIMD (2016) quartile** |  |  |  |
| *1* | 33908 (37.58%) | 2365 (48.16%) | 2302 (37.33%) |
| *2* | 16901 (18.73%) | 895 (18.22%) | 1184 (19.2%) |
| *3* | 14855 (16.46%) | 725 (14.76%) | 1048 (17%) |
| *4* | 23003 (25.49%) | 831 (16.92%) | 1526 (24.75%) |
| *Unknown* | 1566 (1.74%) | 95 (1.93%) | 106 (1.72%) |
| **SARS-CoV-2 positivity status** |  |  |  |
| **Previous confirmed SARS-CoV-2 infection status** |  |  |  |
| *No previous infection* | 83000 (91.98%) | 4812 (97.98%) | 5793 (93.95%) |
| *Had previous infection* | 7233 (8.02%) | 99 (2.02%) | 373 (6.05%) |
| **Vaccination status** |  |  |  |
| **Vaccination schedule** |  |  |  |
| *Unvaccinated* | 13017 (14.45%) | 3017 (61.52%) | 1901 (30.88%) |
| *1st dose AstraZeneca* | 19 (0.02%) | <=5 | <=5 |
| *1st dose Pfizer* | 437 (0.49%) | 26 (0.53%) | 45 (0.73%) |
| *1st dose Moderna* | 295 (0.33%) | 17 (0.35%) | 30 (0.49%) |
| *2nd dose AstraZeneca* | 468 (0.52%) | 19 (0.39%) | 45 (0.73%) |
| *2nd dose Pfizer* | 2448 (2.72%) | 71 (1.45%) | 278 (4.52%) |
| *2nd dose Moderna* | 761 (0.84%) | 17 (0.35%) | 90 (1.46%) |
| *2 x AstraZeneca, 1 x Pfizer* | 23641 (26.25%) | 581 (11.85%) | 1130 (18.36%) |
| *2 x Moderna, 1 x Pfizer* | 235 (0.26%) | 10 (0.2%) | 19 (0.31%) |
| *3 x Pfizer* | 21704 (24.1%) | 491 (10.01%) | 1193 (19.38%) |
| *2 x AstraZeneca, 1 x Moderna* | 15159 (16.83%) | 394 (8.03%) | 743 (12.07%) |
| *2 x Pfizer, 1 x Moderna* | 11581 (12.86%) | 254 (5.18%) | 655 (10.64%) |

|  |  |  |  |  |  |  |  |  |  |
| --- | --- | --- | --- | --- | --- | --- | --- | --- | --- |
| 3 x Moderna | 296 (0.33%) |  |  | <=5 |  |  | 25 (0.41%) |  |  |
| Time (days) since most recent dose by variant and dose number |  |  |  |  |  |  |  |  |  |
|  | Negative |  |  | Delta |  |  | Omicron |  |  |
|  | 1 | 2 | 3 | 1 | 2 | 3 | 1 | 2 | 3 |
| Minimum | 14.00 | 14.00 | 14.00 | 14.00 | 14.00 | 14.00 | 14.00 | 14.00 | 14.0 |
| 1st Quartile | 22.00 | 34.00 | 25.00 | 20.00 | 33.50 | 18.00 | 24.00 | 38.00 | 25.0 |
| Median | 32.00 | 49.00 | 41.00 | 34.00 | 51.00 | 30.00 | 34.00 | 56.00 | 39.0 |
| Mean | 31.15 | 48.32 | 42.04 | 30.29 | 47.53 | 35.44 | 32.71 | 53.15 | 41.5 |
| 3rd Quartile | 39.00 | 65.00 | 58.00 | 37.00 | 65.50 | 50.00 | 40.00 | 69.00 | 58.0 |
| Maximum | 49.00 | 77.00 | 77.00 | 49.00 | 77.00 | 77.00 | 49.00 | 77.00 | 77.0 |

## The COVID-19 Genomics UK (COG-UK) consortium - June 2021 V.1

**Funding acquisition, Leadership and supervision, Metadata curation, Project administration, Samples and logistics, Sequencing and analysis, Software and analysis tools, and Visualisation:**

Dr Samuel C Robson ^13, 84^

**Funding acquisition, Leadership and supervision, Metadata curation, Project administration, Samples and logistics, Sequencing and analysis, and Software and analysis tools:**

Dr Thomas R Connor ^11, 74^ and Prof Nicholas J Loman ^43^

**Leadership and supervision, Metadata curation, Project administration, Samples and logistics, Sequencing and analysis, Software and analysis tools, and Visualisation:**

Dr Tanya Golubchik ^5^

**Funding acquisition, Leadership and supervision, Metadata curation, Samples and logistics, Sequencing and analysis, and Visualisation:**

Dr Rocio T Martinez Nunez ^46^

**Funding acquisition, Leadership and supervision, Project administration, Samples and logistics, Sequencing and analysis, and Software and analysis tools:**

Dr David Bonsall ^5^

**Funding acquisition, Leadership and supervision, Project administration, Sequencing and analysis, Software and analysis tools, and Visualisation:**

Prof Andrew Rambaut ^104^

**Funding acquisition, Metadata curation, Project administration, Samples and logistics, Sequencing and analysis, and Software and analysis tools:**

Dr Luke B Snell ^12^

**Leadership and supervision, Metadata curation, Project administration, Samples and logistics, Software and analysis tools, and Visualisation:**

Rich Livett ^116^

**Funding acquisition, Leadership and supervision, Metadata curation, Project administration, and Samples and logistics:**

Dr Catherine Ludden ^20, 70^

**Funding acquisition, Leadership and supervision, Metadata curation, Samples and logistics, and Sequencing and analysis:**

Dr Sally Corden ^74^ and Dr Eleni Nastouli ^96, 95, 30^

**Funding acquisition, Leadership and supervision, Metadata curation, Sequencing and analysis, and Software and analysis tools:**

Dr Gaia Nebbia ^12^

**Funding acquisition, Leadership and supervision, Project administration, Samples and logistics, and Sequencing and analysis:**

Ian Johnston ^116^

**Leadership and supervision, Metadata curation, Project administration, Samples and logistics, and Sequencing and analysis:**

Prof Katrina Lythgoe ^5^, Dr M. Estee Torok ^19, 20^ and Prof Ian G Goodfellow ^24^

**Leadership and supervision, Metadata curation, Project administration, Samples and logistics, and Visualisation:**

Dr Jacqui A Prieto ^97, 82^ and Dr Kordo Saeed ^97, 83^

**Leadership and supervision, Metadata curation, Project administration, Sequencing and analysis, and Software and analysis tools:**

Dr David K Jackson ^116^

**Leadership and supervision, Metadata curation, Samples and logistics, Sequencing and analysis, and Visualisation:**

Dr Catherine Houlihan ^96, 94^

**Leadership and supervision, Metadata curation, Sequencing and analysis, Software and analysis tools, and Visualisation:**

Dr Dan Frampton ^94, 95^

**Metadata curation, Project administration, Samples and logistics, Sequencing and analysis, and Software and analysis tools:**

Dr William L Hamilton ^19^ and Dr Adam A Witney ^41^

**Funding acquisition, Samples and logistics, Sequencing and analysis, and Visualisation:**

Dr Giselda Bucca ^101^

**Funding acquisition, Leadership and supervision, Metadata curation, and Project administration:**

Dr Cassie F Pope ^40, 41^

**Funding acquisition, Leadership and supervision, Metadata curation, and Samples and logistics:**

Dr Catherine Moore ^74^

**Funding acquisition, Leadership and supervision, Metadata curation, and Sequencing and analysis:**

Prof Emma C Thomson ^53^

**Funding acquisition, Leadership and supervision, Project administration, and Samples and logistics:**

Dr Ewan M Harrison ^116, 102^

**Funding acquisition, Leadership and supervision, Sequencing and analysis, and Visualisation:**

Prof Colin P Smith ^101^

**Leadership and supervision, Metadata curation, Project administration, and Sequencing and analysis:**

Fiona Rogan ^77^

**Leadership and supervision, Metadata curation, Project administration, and Samples and logistics:**

Shaun M Beckwith ^6^, Abigail Murray ^6^, Dawn Singleton ^6^, Dr Kirstine Eastick ^37^, Dr Liz A Sheridan ^98^, Paul Randell ^99^, Dr Leigh M Jackson ^105^, Dr Cristina V Ariani ^116^ and Dr Sónia Gonçalves ^116^

**Leadership and supervision, Metadata curation, Samples and logistics, and Sequencing and analysis:**

Dr Derek J Fairley ^3, 77^, Prof Matthew W Loose ^18^ and Joanne Watkins ^74^

**Leadership and supervision, Metadata curation, Samples and logistics, and Visualisation:**

Dr Samuel Moses ^25, 106^

**Leadership and supervision, Metadata curation, Sequencing and analysis, and Software and analysis tools:**

Dr Sam Nicholls ^43^, Dr Matthew Bull ^74^ and Dr Roberto Amato ^116^

**Leadership and supervision, Project administration, Samples and logistics, and Sequencing and analysis:**

Prof Darren L Smith ^36, 65, 66^

**Leadership and supervision, Sequencing and analysis, Software and analysis tools, and Visualisation:**

Prof David M Aanensen ^14, 116^ and Dr Jeffrey C Barrett ^116^

**Metadata curation, Project administration, Samples and logistics, and Sequencing and analysis:**

Dr Dinesh Aggarwal ^20, 116, 70^, Dr James G Shepherd ^53^, Dr Martin D Curran ^71^ and Dr Surendra Parmar71

**Metadata curation, Project administration, Sequencing and analysis, and Software and analysis tools:**

Dr Matthew D Parker ^109^

**Metadata curation, Samples and logistics, Sequencing and analysis, and Software and analysis tools:**

Dr Catryn Williams ^74^

**Metadata curation, Samples and logistics, Sequencing and analysis, and Visualisation:**

Dr Sharon Glaysher ^68^

**Metadata curation, Sequencing and analysis, Software and analysis tools, and Visualisation:**

Dr Anthony P Underwood ^14, 116^, Dr Matthew Bashton ^36, 65^, Dr Nicole Pacchiarini ^74^, Dr Katie F Loveson ^84^ and Matthew Byott ^95, 96^

**Project administration, Sequencing and analysis, Software and analysis tools, and Visualisation:**

Dr Alessandro M Carabelli ^20^

**Funding acquisition, Leadership and supervision, and Metadata curation:**

Dr Kate E Templeton ^56, 104^

**Funding acquisition, Leadership and supervision, and Project administration:**

Dr Thushan I de Silva ^109^, Dr Dennis Wang ^109^, Dr Cordelia F Langford ^116^ and John Sillitoe ^116^

**Funding acquisition, Leadership and supervision, and Samples and logistics:**

Prof Rory N Gunson ^55^

**Funding acquisition, Leadership and supervision, and Sequencing and analysis:**

Dr Simon Cottrell ^74^, Dr Justin O’Grady ^75, 103^ and Prof Dominic Kwiatkowski ^116, 108^

**Leadership and supervision, Metadata curation, and Project administration:**

Dr Patrick J Lillie ^37^

**Leadership and supervision, Metadata curation, and Samples and logistics:**

Dr Nicholas Cortes ^33^, Dr Nathan Moore ^33^, Dr Claire Thomas ^33^, Phillipa J Burns ^37^, Dr Tabitha W Mahungu ^80^ and Steven Liggett ^86^

**Leadership and supervision, Metadata curation, and Sequencing and analysis:**

Angela H Beckett ^13, 81^ and Prof Matthew TG Holden ^73^

**Leadership and supervision, Project administration, and Samples and logistics:**

Dr Lisa J Levett ^34^, Dr Husam Osman ^70, 35^ and Dr Mohammed O Hassan-Ibrahim ^99^

**Leadership and supervision, Project administration, and Sequencing and analysis:**

Dr David A Simpson ^77^

**Leadership and supervision, Samples and logistics, and Sequencing and analysis:**

Dr Meera Chand ^72^, Prof Ravi K Gupta ^102^, Prof Alistair C Darby ^107^ and Prof Steve Paterson ^107^

**Leadership and supervision, Sequencing and analysis, and Software and analysis tools:**

Prof Oliver G Pybus ^23^, Dr Erik M Volz ^39^, Prof Daniela de Angelis ^52^, Prof David L Robertson ^53^, Dr Andrew J Page ^75^ and Dr Inigo Martincorena ^116^

**Leadership and supervision, Sequencing and analysis, and Visualisation:**

Dr Louise Aigrain ^116^ and Dr Andrew R Bassett ^116^

**Metadata curation, Project administration, and Samples and logistics:**

Dr Nick Wong ^50^, Dr Yusri Taha ^89^, Michelle J Erkiert ^99^ and Dr Michael H Spencer Chapman ^116, 102^

**Metadata curation, Project administration, and Sequencing and analysis:**

Dr Rebecca Dewar ^56^ and Martin P McHugh ^56, 111^

**Metadata curation, Project administration, and Software and analysis tools:**

Siddharth Mookerjee ^38, 57^

**Metadata curation, Project administration, and Visualisation:**

Stephen Aplin ^97^, Matthew Harvey ^97^, Thea Sass ^97^, Dr Helen Umpleby ^97^ and Helen Wheeler ^97^

**Metadata curation, Samples and logistics, and Sequencing and analysis:**

Dr James P McKenna ^3^, Dr Ben Warne ^9^, Joshua F Taylor ^22^, Yasmin Chaudhry ^24^, Rhys Izuagbe ^24^, Dr Aminu S Jahun ^24^, Dr Gregory R Young ^36, 65^, Dr Claire McMurray ^43^, Dr Clare M McCann ^65, 66^, Dr Andrew Nelson ^65, 66^ and Scott Elliott ^68^

**Metadata curation, Samples and logistics, and Visualisation:**

Hannah Lowe ^25^

**Metadata curation, Sequencing and analysis, and Software and analysis tools:**

Dr Anna Price ^11^, Matthew R Crown ^65^, Dr Sara Rey ^74^, Dr Sunando Roy ^96^ and Dr Ben Temperton ^105^

**Metadata curation, Sequencing and analysis, and Visualisation:**

Dr Sharif Shaaban ^73^ and Dr Andrew R Hesketh ^101^

**Project administration, Samples and logistics, and Sequencing and analysis:**

Dr Kenneth G Laing ^41^, Dr Irene M Monahan ^41^ and Dr Judith Heaney ^95, 96, 34^

**Project administration, Samples and logistics, and Visualisation:**

Dr Emanuela Pelosi ^97^, Siona Silviera ^97^ and Dr Eleri Wilson-Davies ^97^

**Samples and logistics, Software and analysis tools, and Visualisation:**

Dr Helen Fryer ^5^

**Sequencing and analysis, Software and analysis tools, and Visualization:**

Dr Helen Adams ^4^, Dr Louis du Plessis ^23^, Dr Rob Johnson ^39^, Dr William T Harvey ^53, 42^, Dr Joseph Hughes ^53^, Dr Richard J Orton ^53^, Dr Lewis G Spurgin ^59^, Dr Yann Bourgeois ^81^, Dr Chris Ruis ^102^, Áine O’Toole ^104^, Marina Gourtovaia ^116^ and Dr Theo Sanderson ^116^

**Funding acquisition, and Leadership and supervision:**

Dr Christophe Fraser ^5^, Dr Jonathan Edgeworth ^12^, Prof Judith Breuer ^96, 29^, Dr Stephen L Michell ^105^ and Prof John A Todd ^115^

**Funding acquisition, and Project administration:**

Michaela John ^10^ and Dr David Buck ^115^

**Leadership and supervision, and Metadata curation:**

Dr Kavitha Gajee ^37^ and Dr Gemma L Kay ^75^

**Leadership and supervision, and Project administration:**

Prof Sharon J Peacock ^20, 70^ and David Heyburn ^74^

**Leadership and supervision, and Samples and logistics:**

Katie Kitchman ^37^, Prof Alan McNally ^43, 93^, David T Pritchard ^50^, Dr Samir Dervisevic ^58^, Dr Peter Muir^70^, Dr Esther Robinson ^70, 35^, Dr Barry B Vipond ^70^, Newara A Ramadan ^78^, Dr Christopher Jeanes ^90^, Danni Weldon ^116^, Jana Catalan ^118^ and Neil Jones ^118^

**Leadership and supervision, and Sequencing and analysis:**

Dr Ana da Silva Filipe ^53^, Dr Chris Williams ^74^, Marc Fuchs ^77^, Dr Julia Miskelly ^77^, Dr Aaron R Jeffries ^105^, Karen Oliver ^116^ and Dr Naomi R Park ^116^

**Metadata curation, and Samples and logistics:**

Amy Ash ^1^, Cherian Koshy ^1^, Magdalena Barrow ^7^, Dr Sarah L Buchan ^7^, Dr Anna Mantzouratou ^7^, Dr Gemma Clark ^15^, Dr Christopher W Holmes ^16^, Sharon Campbell ^17^, Thomas Davis ^21^, Ngee Keong Tan ^22^, Dr Julianne R Brown ^29^, Dr Kathryn A Harris ^29, 2^, Stephen P Kidd ^33^, Dr Paul R Grant ^34^, Dr Li Xu-McCrae ^35^, Dr Alison Cox ^38, 63^, Pinglawathee Madona ^38, 63^, Dr Marcus Pond ^38, 63^, Dr Paul A Randell ^38, 63^, Karen T Withell ^48^, Cheryl Williams ^51^, Dr Clive Graham ^60^, Rebecca Denton-Smith ^62^, Emma Swindells ^62^, Robyn Turnbull ^62^, Dr Tim J Sloan ^67^, Dr Andrew Bosworth ^70, 35^, Stephanie Hutchings ^70^, Hannah M Pymont ^70^, Dr Anna Casey ^76^, Dr Liz Ratcliffe ^76^, Dr Christopher R Jones ^79, 105^, Dr Bridget A Knight ^79, 105^, Dr Tanzina Haque ^80^, Dr Jennifer Hart ^80^, Dr Dianne Irish-Tavares ^80^, Eric Witele ^80^, Craig Mower ^86^, Louisa K Watson ^86^, Jennifer Collins ^89^, Gary Eltringham ^89^, Dorian Crudgington ^98^, Ben Macklin ^98^, Prof Miren Iturriza-Gomara ^107^, Dr Anita O Lucaci ^107^ and Dr Patrick C McClure ^113^

**Metadata curation, and Sequencing and analysis:**

Matthew Carlile ^18^, Dr Nadine Holmes ^18^, Dr Christopher Moore ^18^, Dr Nathaniel Storey ^29^, Dr Stefan Rooke ^73^, Dr Gonzalo Yebra ^73^, Dr Noel Craine ^74^, Malorie Perry ^74^, Dr Nabil-Fareed Alikhan ^75^, Dr Stephen Bridgett ^77^, Kate F Cook ^84^, Christopher Fearn ^84^, Dr Salman Goudarzi ^84^, Prof Ronan A Lyons ^88^, Dr Thomas Williams ^104^, Dr Sam T Haldenby ^107^, Jillian Durham ^116^ and Dr Steven Leonard ^116^

**Metadata curation, and Software and analysis tools:**

Robert M Davies ^116^

**Project administration, and Samples and logistics:**

Dr Rahul Batra ^12^, Beth Blane ^20^, Dr Moira J Spyer ^30, 95, 96^, Perminder Smith ^32, 112^, Mehmet Yavus ^85, 109^, Dr Rachel J Williams ^96^, Dr Adhyana IK Mahanama ^97^, Dr Buddhini Samaraweera ^97^, Sophia T Girgis ^102^, Samantha E Hansford ^109^, Dr Angie Green ^115^, Dr Charlotte Beaver ^116^, Katherine L Bellis ^116, 102^, Matthew J Dorman ^116^, Sally Kay ^116^, Liam Prestwood ^116^ and Dr Shavanthi Rajatileka ^116^

**Project administration, and Sequencing and analysis:**

Dr Joshua Quick ^43^

**Project administration, and Software and analysis tools:**

Radoslaw Poplawski ^43^

**Samples and logistics, and Sequencing and analysis:**

Dr Nicola Reynolds ^8^, Andrew Mack ^11^, Dr Arthur Morriss ^11^, Thomas Whalley ^11^, Bindi Patel ^12^, Dr Iliana Georgana ^24^, Dr Myra Hosmillo ^24^, Malte L Pinckert ^24^, Dr Joanne Stockton ^43^, Dr John H Henderson ^65^, Amy Hollis ^65^, Dr William Stanley ^65^, Dr Wen C Yew ^65^, Dr Richard Myers ^72^, Dr Alicia Thornton ^72^, Alexander Adams ^74^, Tara Annett ^74^, Dr Hibo Asad ^74^, Alec Birchley ^74^, Jason Coombes ^74^, Johnathan M Evans ^74^, Laia Fina ^74^, Bree Gatica-Wilcox ^74^, Lauren Gilbert ^74^, Lee Graham ^74^, Jessica Hey ^74^, Ember Hilvers ^74^, Sophie Jones ^74^, Hannah Jones ^74^, Sara Kumziene-Summerhayes ^74^, Dr Caoimhe McKerr ^74^, Jessica Powell ^74^, Georgia Pugh ^74^, Sarah Taylor ^74^, Alexander J Trotter ^75^, Charlotte A Williams ^96^, Leanne M Kermack ^102^, Benjamin H Foulkes ^109^, Marta Gallis ^109^, Hailey R Hornsby ^109^, Stavroula F Louka ^109^, Dr Manoj Pohare ^109^, Paige Wolverson ^109^, Peijun Zhang ^109^, George MacIntyre-Cockett ^115^, Amy Trebes ^115^, Dr Robin J Moll ^116^, Lynne Ferguson ^117^, Dr Emily J Goldstein ^117^, Dr Alasdair Maclean ^117^ and Dr Rachael Tomb ^117^

**Samples and logistics, and Software and analysis tools:**

Dr Igor Starinskij ^53^

**Sequencing and analysis, and Software and analysis tools:**

Laura Thomson ^5^, Joel Southgate ^11, 74^, Dr Moritz UG Kraemer ^23^, Dr Jayna Raghwani ^23^, Dr Alex E Zarebski ^23^, Olivia Boyd ^39^, Lily Geidelberg ^39^, Dr Chris J Illingworth ^52^, Dr Chris Jackson ^52^, Dr David Pascall ^52^, Dr Sreenu Vattipally ^53^, Timothy M Freeman ^109^, Dr Sharon N Hsu ^109^, Dr Benjamin B Lindsey^109^, Dr Keith James ^116^, Kevin Lewis ^116^, Gerry Tonkin-Hill ^116^ and Dr Jaime M Tovar-Corona ^116^

**Sequencing and analysis, and Visualisation:**

MacGregor Cox ^20^

**Software and analysis tools, and Visualisation:**

Dr Khalil Abudahab ^14, 116^, Mirko Menegazzo ^14^, Ben EW Taylor MEng ^14, 116^, Dr Corin A Yeats ^14^, Afrida Mukaddas ^53^, Derek W Wright ^53^, Dr Leonardo de Oliveira Martins ^75^, Dr Rachel Colquhoun ^104^, Verity Hill ^104^, Dr Ben Jackson ^104^, Dr JT McCrone ^104^, Dr Nathan Medd ^104^, Dr Emily Scher ^104^ and Jon-Paul Keatley ^116^

**Leadership and supervision:**

Dr Tanya Curran ^3^, Dr Sian Morgan ^10^, Prof Patrick Maxwell ^20^, Prof Ken Smith ^20^, Dr Sahar Eldirdiri ^21^, Anita Kenyon ^21^, Prof Alison H Holmes ^38, 57^, Dr James R Price ^38, 57^, Dr Tim Wyatt ^69^, Dr Alison E Mather ^75^, Dr Timofey Skvortsov ^77^ and Prof John A Hartley ^96^

**Metadata curation:**

Prof Martyn Guest ^11^, Dr Christine Kitchen ^11^, Dr Ian Merrick ^11^, Robert Munn ^11^, Dr Beatrice Bertolusso ^33^, Dr Jessica Lynch ^33^, Dr Gabrielle Vernet ^33^, Stuart Kirk ^34^, Dr Elizabeth Wastnedge ^56^, Dr Rachael Stanley ^58^, Giles Idle ^64^, Dr Declan T Bradley ^69, 77^, Dr Jennifer Poyner ^79^ and Matilde Mori ^110^

**Project administration:**

Owen Jones ^11^, Victoria Wright ^18^, Ellena Brooks ^20^, Carol M Churcher ^20^, Mireille Fragakis ^20^, Dr Katerina Galai ^20, 70^, Dr Andrew Jermy ^20^, Sarah Judges ^20^, Georgina M McManus ^20^, Kim S Smith ^20^, Dr Elaine Westwick ^20^, Dr Stephen W Attwood ^23^, Dr Frances Bolt ^38, 57^, Dr Alisha Davies ^74^, Elen De Lacy ^74^, Fatima Downing ^74^, Sue Edwards ^74^, Lizzie Meadows ^75^, Sarah Jeremiah ^97^, Dr Nikki Smith ^109^ and Luke Foulser ^116^

**Samples and logistics:**

Dr Themoula Charalampous ^12, 46^, Amita Patel ^12^, Dr Louise Berry ^15^, Dr Tim Boswell ^15^, Dr Vicki M Fleming ^15^, Dr Hannah C Howson-Wells ^15^, Dr Amelia Joseph ^15^, Manjinder Khakh ^15^, Dr Michelle M Lister ^15^, Paul W Bird ^16^, Karlie Fallon ^16^, Thomas Helmer ^16^, Dr Claire L McMurray ^16^, Mina Odedra ^16^, Jessica Shaw ^16^, Dr Julian W Tang ^16^, Nicholas J Willford ^16^, Victoria Blakey ^17^, Dr Veena Raviprakash ^17^, Nicola Sheriff ^17^, Lesley-Anne Williams ^17^, Theresa Feltwell ^20^, Dr Luke Bedford ^26^, Dr James S Cargill ^27^, Warwick Hughes ^27^, Dr Jonathan Moore ^28^, Susanne Stonehouse ^28^, Laura Atkinson ^29^, Jack CD Lee ^29^, Dr Divya Shah ^29^, Adela Alcolea-Medina ^32, 112^, Natasha Ohemeng-Kumi ^32, 112^, John Ramble ^32, 112^, Jasveen Sehmi ^32, 112^, Dr Rebecca Williams ^33^, Wendy Chatterton ^34^, Monika Pusok ^34^, William Everson ^37^, Anibolina Castigador ^44^, Emily Macnaughton ^44^, Dr Kate El Bouzidi ^45^, Dr Temi Lampejo ^45^, Dr Malur Sudhanva ^45^, Cassie Breen ^47^, Dr Graciela Sluga ^48^, Dr Shazaad SY Ahmad ^49, 70^, Dr Ryan P George ^49^, Dr Nicholas W Machin ^49, 70^, Debbie Binns ^50^, Victoria James ^50^, Dr Rachel Blacow ^55^, Dr Lindsay Coupland ^58^, Dr Louise Smith ^59^, Dr Edward Barton ^60^, Debra Padgett ^60^, Garren Scott ^60^, Dr Aidan Cross ^61^, Dr Mariyam Mirfenderesky ^61^, Jane Greenaway ^62^, Kevin Cole ^64^, Phillip Clarke ^67^, Nichola Duckworth ^67^, Sarah Walsh ^67^, Kelly Bicknell ^68^, Robert Impey ^68^, Dr Sarah Wyllie ^68^, Richard Hopes ^70^, Dr Chloe Bishop ^72^, Dr Vicki Chalker ^72^, Dr Ian Harrison ^72^, Laura Gifford ^74^, Dr Zoltan Molnar ^77^, Dr Cressida Auckland ^79^, Dr Cariad Evans ^85, 109^, Dr Kate Johnson ^85, 109^, Dr David G Partridge ^85, 109^, Dr Mohammad Raza ^85, 109^, Paul Baker ^86^, Prof Stephen Bonner ^86^, Sarah Essex ^86^, Leanne J Murray ^86^, Andrew I Lawton ^87^, Dr Shirelle Burton-Fanning ^89^, Dr Brendan AI Payne ^89^, Dr Sheila Waugh ^89^, Andrea N Gomes ^91^, Maimuna Kimuli ^91^, Darren R Murray ^91^, Paula Ashfield ^92^, Dr Donald Dobie ^92^, Dr Fiona Ashford ^93^, Dr Angus Best ^93^, Dr Liam Crawford ^93^, Dr Nicola Cumley ^93^, Dr Megan Mayhew ^93^, Dr Oliver Megram ^93^, Dr Jeremy Mirza ^93^, Dr Emma Moles-Garcia ^93^, Dr Benita Percival ^93^, Megan Driscoll ^96^, Leah Ensell ^96^, Dr Helen L Lowe ^96^, Laurentiu Maftei ^96^, Matteo Mondani ^96^, Nicola J Chaloner ^99^, Benjamin J Cogger^99^, Lisa J Easton ^99^, Hannah Huckson ^99^, Jonathan Lewis ^99^, Sarah Lowdon ^99^, Cassandra S Malone ^99^, Florence Munemo ^99^, Manasa Mutingwende ^99^, Roberto Nicodemi ^99^, Olga Podplomyk ^99^, Thomas Somassa ^99^, Dr Andrew Beggs ^100^, Dr Alex Richter ^100^, Claire Cormie ^102^, Joana Dias ^102^, Sally Forrest ^102^, Dr Ellen E Higginson ^102^, Mailis Maes ^102^, Jamie Young ^102^, Dr Rose K Davidson ^103^, Kathryn A Jackson ^107^, Dr Lance Turtle ^107^, Dr Alexander J Keeley ^109^, Prof Jonathan Ball ^113^, Timothy Byaruhanga ^113^, Dr Joseph G Chappell ^113^, Jayasree Dey ^113^, Jack D Hill ^113^, Emily J Park ^113^, Arezou Fanaie ^114^, Rachel A Hilson ^114^, Geraldine Yaze ^114^ and Stephanie Lo ^116^

**Sequencing and analysis:**

Safiah Afifi ^10^, Robert Beer ^10^, Joshua Maksimovic ^10^, Kathryn McCluggage ^10^, Karla Spellman ^10^, Catherine Bresner ^11^, William Fuller ^11^, Dr Angela Marchbank ^11^, Trudy Workman ^11^, Dr Ekaterina Shelest ^13, 81^, Dr Johnny Debebe ^18^, Dr Fei Sang ^18^, Dr Marina Escalera Zamudio ^23^, Dr Sarah Francois ^23^, Bernardo Gutierrez ^23^, Dr Tetyana I Vasylyeva ^23^, Dr Flavia Flaviani ^31^, Dr Manon Ragonnet-Cronin ^39^, Dr Katherine L Smollett ^42^, Alice Broos ^53^, Daniel Mair ^53^, Jenna Nichols ^53^, Dr Kyriaki Nomikou ^53^, Dr Lily Tong ^53^, Ioulia Tsatsani ^53^, Prof Sarah O’Brien ^54^, Prof Steven Rushton ^54^, Dr Roy Sanderson ^54^, Dr Jon Perkins ^55^, Seb Cotton ^56^, Abbie Gallagher ^56^, Dr Elias Allara ^70, 102^, Clare Pearson ^70, 102^, Dr David Bibby ^72^, Dr Gavin Dabrera ^72^, Dr Nicholas Ellaby ^72^, Dr Eileen Gallagher ^72^, Dr Jonathan Hubb ^72^, Dr Angie Lackenby ^72^, Dr David Lee ^72^, Nikos Manesis ^72^, Dr Tamyo Mbisa ^72^, Dr Steven Platt ^72^, Katherine A Twohig ^72^, Dr Mari Morgan ^74^, Alp Aydin ^75^, David J Baker ^75^, Dr Ebenezer Foster-Nyarko ^75^, Dr Sophie J Prosolek ^75^, Steven Rudder ^75^, Chris Baxter ^77^, Sílvia F Carvalho ^77^, Dr Deborah Lavin ^77^, Dr Arun Mariappan ^77^, Dr Clara Radulescu ^77^, Dr Aditi Singh ^77^, Miao Tang ^77^, Helen Morcrette ^79^, Nadua Bayzid ^96^, Marius Cotic ^96^, Dr Carlos E Balcazar ^104^, Dr Michael D Gallagher ^104^, Dr Daniel Maloney ^104^, Thomas D Stanton ^104^, Dr Kathleen A Williamson ^104^, Dr Robin Manley ^105^, Michelle L Michelsen ^105^, Dr Christine M Sambles ^105^, Dr David J Studholme ^105^, Joanna Warwick-Dugdale ^105^, Richard Eccles ^107^, Matthew Gemmell ^107^, Dr Richard Gregory ^107^, Dr Margaret Hughes ^107^, Charlotte Nelson ^107^, Dr Lucille Rainbow ^107^, Dr Edith E Vamos ^107^, Hermione J Webster ^107^, Dr Mark Whitehead ^107^, Claudia Wierzbicki ^107^, Dr Adrienn Angyal ^109^, Dr Luke R Green ^109^, Dr Max Whiteley ^109^, Emma Betteridge ^116^, Dr Iraad F Bronner ^116^, Ben W Farr ^116^, Scott Goodwin ^116^, Dr Stefanie V Lensing ^116^, Shane A McCarthy ^116, 102^, Dr Michael A Quail ^116^, Diana Rajan ^116^, Dr Nicholas M Redshaw ^116^, Carol Scott ^116^, Lesley Shirley ^116^ and Scott AJ Thurston ^116^

**Software and analysis tools:**

Dr Will Rowe ^43^, Amy Gaskin ^74^, Dr Thanh Le-Viet ^75^, James Bonfield ^116^, Jennifier Liddle ^116^ and Andrew Whitwham ^116^

**1** Barking, Havering and Redbridge University Hospitals NHS Trust, **2** Barts Health NHS Trust, **3** Belfast Health & Social Care Trust, **4** Betsi Cadwaladr University Health Board, **5** Big Data Institute, Nuffield Department of Medicine, University of Oxford, **6** Blackpool Teaching Hospitals NHS Foundation Trust, **7** Bournemouth University, **8** Cambridge Stem Cell Institute, University of Cambridge, **9** Cambridge University Hospitals NHS Foundation Trust, **10** Cardiff and Vale University Health Board, **11** Cardiff University, **12** Centre for Clinical Infection and Diagnostics Research, Department of Infectious Diseases, Guy’s and St Thomas’ NHS Foundation Trust, **13** Centre for Enzyme Innovation, University of Portsmouth, **14** Centre for Genomic Pathogen Surveillance, University of Oxford, **15** Clinical Microbiology Department, Queens Medical Centre, Nottingham University Hospitals NHS Trust, **16** Clinical Microbiology, University Hospitals of Leicester NHS Trust, **17** County Durham and Darlington NHS Foundation Trust, **18** Deep Seq, School of Life Sciences, Queens Medical Centre, University of Nottingham, **19** Department of Infectious Diseases and Microbiology, Cambridge University Hospitals NHS Foundation Trust, **20** Department of Medicine, University of Cambridge, **21** Department of Microbiology, Kettering General Hospital, **22** Department of Microbiology, South West London Pathology, **23** Department of Zoology, University of Oxford, **24** Division of Virology, Department of Pathology, University of Cambridge, **25** East Kent Hospitals University NHS Foundation Trust, **26** East Suffolk and North Essex NHS Foundation Trust, **27** East Sussex Healthcare NHS Trust**, 28** Gateshead Health NHS Foundation Trust, **29** Great Ormond Street Hospital for Children NHS Foundation Trust, **30** Great Ormond Street Institute of Child Health (GOS ICH), University College London (UCL), **31** Guy’s and St. Thomas’ Biomedical Research Centre, **32** Guy’s and St. Thomas’ NHS Foundation Trust, **33** Hampshire Hospitals NHS Foundation Trust, **34** Health Services Laboratories, **35** Heartlands Hospital, Birmingham, **36** Hub for Biotechnology in the Built Environment, Northumbria University, **37** Hull University Teaching Hospitals NHS Trust, **38** Imperial College Healthcare NHS Trust, **39** Imperial College London, **40** Infection Care Group, St George’s University Hospitals NHS Foundation Trust, **41** Institute for Infection and Immunity, St George’s University of London, **42** Institute of Biodiversity, Animal Health & Comparative Medicine, **43** Institute of Microbiology and Infection, University of Birmingham, **44** Isle of Wight NHS Trust, **45** King’s College Hospital NHS Foundation Trust, **46** King’s College London, **47** Liverpool Clinical Laboratories, **48** Maidstone and Tunbridge Wells NHS Trust, **49** Manchester University NHS Foundation Trust, **50** Microbiology Department, Buckinghamshire Healthcare NHS Trust, **51** Microbiology, Royal Oldham Hospital, **52** MRC Biostatistics Unit, University of Cambridge, **53** MRC-University of Glasgow Centre for Virus Research, **54** Newcastle University, **55** NHS Greater Glasgow and Clyde, **56** NHS Lothian, **57** NIHR Health Protection Research Unit in HCAI and AMR, Imperial College London, **58** Norfolk and Norwich University Hospitals NHS Foundation Trust, **59** Norfolk County Council, **60** North Cumbria Integrated Care NHS Foundation Trust, **61** North Middlesex University Hospital NHS Trust, **62** North Tees and Hartlepool NHS Foundation Trust, **63** North West London Pathology, **64** Northumbria Healthcare NHS Foundation Trust, **65** Northumbria University, **66** NU-OMICS, Northumbria University, **67** Path Links, Northern Lincolnshire and Goole NHS Foundation Trust, **68** Portsmouth Hospitals University NHS Trust, **69** Public Health Agency, Northern Ireland, **70** Public Health England, **71** Public Health England, Cambridge, **72** Public Health England, Colindale, **73** Public Health Scotland, **74** Public Health Wales, **75** Quadram Institute Bioscience, **76** Queen Elizabeth Hospital, Birmingham, **77** Queen’s University Belfast, **78** Royal Brompton and Harefield Hospitals, **79** Royal Devon and Exeter NHS Foundation Trust, **80** Royal Free London NHS Foundation Trust, **81** School of Biological Sciences, University of Portsmouth, **82** School of Health Sciences, University of Southampton, **83** School of Medicine, University of Southampton, **84** School of Pharmacy & Biomedical Sciences, University of Portsmouth, **85** Sheffield Teaching Hospitals NHS Foundation Trust, **86** South Tees Hospitals NHS Foundation Trust, **87** Southwest Pathology Services, **88** Swansea University, **89** The Newcastle upon Tyne Hospitals NHS Foundation Trust, **90** The Queen Elizabeth Hospital King’s Lynn NHS Foundation Trust, **91** The Royal Marsden NHS Foundation Trust, **92** The Royal Wolverhampton NHS Trust, **93** Turnkey Laboratory, University of Birmingham, **94** University College London Division of Infection and Immunity**, 95** University College London Hospital Advanced Pathogen Diagnostics Unit**, 96** University College London Hospitals NHS Foundation Trust, **97** University Hospital Southampton NHS Foundation Trust, **98** University Hospitals Dorset NHS Foundation Trust, **99** University Hospitals Sussex NHS Foundation Trust, **100** University of Birmingham, **101** University of Brighton, **102** University of Cambridge, **103** University of East Anglia, **104** University of Edinburgh, **105** University of Exeter, **106** University of Kent, **107** University of Liverpool, **108** University of Oxford, **109** University of Sheffield, **110** University of Southampton, **111** University of St Andrews, **112** Viapath, Guy’s and St Thomas’ NHS Foundation Trust, and King’s College Hospital NHS Foundation Trust, **113** Virology, School of Life Sciences, Queens Medical Centre, University of Nottingham, **114** Watford General Hospital, **115** Wellcome Centre for Human Genetics, Nuffield Department of Medicine, University of Oxford, **116** Wellcome Sanger Institute, **117** West of Scotland Specialist Virology Centre, NHS Greater Glasgow and Clyde, **118** Whittington Health NHS Trust

## References

1. WHO SPRP 2021 Mid-term Report - WHO Strategic Action Against COVID 19.

2. Davis, C. et al. Reduced neutralisation of the Delta (B.1.617.2) SARS-CoV-2 variant of concern following vaccination. PLoS Pathog 17, e1010022 (2021).

3. Wall, E.C. et al. Neutralising antibody activity against SARS-CoV-2 VOCs B.1.617.2 and B.1.351 by BNT162b2 vaccination. The Lancet 397, 2331–2333 (2021).

4. Wall, E.C., et al. AZD1222-induced neutralising antibody activity against SARS-CoV-2 Delta VOC. Lancet 398, 207–209 (2021).

5. Lopez Bernal, J., et al. Effectiveness of Covid-19 Vaccines against the B.1.617.2 (Delta) Variant. New England Journal of Medicine 385, 585–594 (2021).

6. Viana, R., et al. Rapid epidemic expansion of the SARS-CoV-2 Omicron variant in southern Africa. medRxiv, 2021.2012.2019.21268028 (2021).

7. Gu, H. et al. Probable Transmission of SARS-CoV-2 Omicron Variant in Quarantine Hotel, Hong Kong, China, November 2021. Emerg Infect Dis 28 (2021).

8. Cameroni, E. et al. Broadly neutralizing antibodies overcome SARS-CoV-2 Omicron antigenic shift. Nature (2021).

9. 9. Andrews, N., et al. Effectiveness of COVID-19 vaccines against the Omicron (B.1.1.529) variant of concern. medRxiv, 2021.2012.2014.21267615 (2021).

10. Aggarwal, A., et al. SARS-CoV-2 Omicron: evasion of potent humoral responses and resistance to clinical immunotherapeutics relative to viral variants of concern. medRxiv, 2021.2012.2014.21267772 (2021).

11. Basile, K., et al. Improved neutralization of the SARS-CoV-2 Omicron variant after Pfizer-BioNTech BNT162b2 COVID-19 vaccine boosting. bioRxiv, 2021.2012.2012.472252 (2021).

12. Ahmed, S.F., Quadeer, A.A. & McKay, M.R. SARS-CoV-2 T cell responses are expected to remain robust against Omicron. bioRxiv, 2021.2012.2012.472315 (2021).

13. Cao, Y. et al. Omicron escapes the majority of existing SARS-CoV-2 neutralizing antibodies. Nature (2021).

14. Cele, S., et al. SARS-CoV-2 Omicron has extensive but incomplete escape of Pfizer BNT162b2 elicited neutralization and requires ACE2 for infection. medRxiv, 2021.2012.2008.21267417 (2021).

15. Dejnirattisai, W. et al. Reduced neutralisation of SARS-CoV-2 omicron B.1.1.529 variant by post-immunisation serum. The Lancet (2021).

16. Doria-Rose, N.A., et al. Booster of mRNA-1273 Vaccine Reduces SARS-CoV-2 Omicron Escape from Neutralizing Antibodies. medRxiv, 2021.2012.2015.21267805 (2021).

17. Garcia-Beltran, W.F. et al. mRNA-based COVID-19 vaccine boosters induce neutralizing immunity against SARS-CoV-2 Omicron variant. Cell (2022).

18. Meng, B., et al. SARS-CoV-2 Omicron spike mediated immune escape, infectivity and cell-cell fusion. bioRxiv, 2021.2012.2017.473248 (2021).

19. Zahradník, J. et al. SARS-CoV-2 variant prediction and antiviral drug design are enabled by RBD in vitro evolution. Nature Microbiology 6, 1188–1198 (2021).

20. Greaney, A.J. et al. Comprehensive mapping of mutations in the SARS-CoV-2 receptor- binding domain that affect recognition by polyclonal human plasma antibodies. Cell host & microbe 29, 463–476 e466 (2021).

21. Burnett, D.L. et al. Immunizations with diverse sarbecovirus receptor-binding domains elicit SARS-CoV-2 neutralizing antibodies against a conserved site of vulnerability. Immunity 54, 2908–2921.e2906 (2021).

22. Pinto, D. et al. Cross-neutralization of SARS-CoV-2 by a human monoclonal SARS-CoV antibody. Nature 583, 290–295 (2020).

23. Rouet, R. et al. Potent SARS-CoV-2 binding and neutralization through maturation of iconic SARS-CoV-1 antibodies. MAbs 13, 1922134 (2021).

24. Peacock, T.P. et al. The furin cleavage site in the SARS-CoV-2 spike protein is required for transmission in ferrets. Nature Microbiology 6, 899–909 (2021).

25. Meng, B. et al. Recurrent emergence of SARS-CoV-2 spike deletion H69/V70 and its role in the Alpha variant B.1.1.7. Cell Reports 35, 109292 (2021).

26. McCallum, M. et al. N-terminal domain antigenic mapping reveals a site of vulnerability for SARS-CoV-2. Cell 184, 2332–2347.e2316 (2021).

27. McCarthy, K.R. et al. Recurrent deletions in the SARS-CoV-2 spike glycoprotein drive antibody escape. *Science*, eabf6950 (2021).

28. Greaney, A.J. et al. Mapping mutations to the SARS-CoV-2 RBD that escape binding by different classes of antibodies. Nat. Commun. 12, 4196 (2021).

29. Woo, H. et al. Developing a Fully Glycosylated Full-Length SARS-CoV-2 Spike Protein Model in a Viral Membrane. J Phys Chem B 124, 7128–7137 (2020).

30. Wrapp, D. et al. Cryo-EM structure of the 2019-nCoV spike in the prefusion conformation. Science 367, 1260–1263 (2020).

31. Sweredoski, M.J. & Baldi, P. PEPITO: improved discontinuous B-cell epitope prediction using multiple distance thresholds and half sphere exposure. Bioinformatics 24, 1459–1460 (2008).

32. McCallum, M., et al. SARS-CoV-2 immune evasion by variant B.1.427/B.1.429. bioRxiv, 2021.2003.2031.437925 (2021).

33. Starr, T.N. et al. Prospective mapping of viral mutations that escape antibodies used to treat COVID-19. Science 371, 850–854 (2021).

34. Weisblum, Y. et al. Escape from neutralizing antibodies by SARS-CoV-2 spike protein variants. Elife 9 (2020).

35. Liu, Z. et al. Identification of SARS-CoV-2 spike mutations that attenuate monoclonal and serum antibody neutralization. Cell host & microbe (2021).

36. Wang, Z., et al. mRNA vaccine-elicited antibodies to SARS-CoV-2 and circulating variants. bioRxiv, 2021.2001.2015.426911 (2021).

37. Lan, J. et al. Structure of the SARS-CoV-2 spike receptor-binding domain bound to the ACE2 receptor. Nature 581, 215–220 (2020).

38. Starr, T.N. et al. Deep Mutational Scanning of SARS-CoV-2 Receptor Binding Domain Reveals Constraints on Folding and ACE2 Binding. Cell 182, 1295–1310 e1220 (2020).

39. Cromer, D. et al. Neutralising antibody titres as predictors of protection against SARS-CoV-2 variants and the impact of boosting: a meta-analysis. The Lancet Microbe (2021).

40. Gilbert, P.B., et al. Immune Correlates Analysis of the mRNA-1273 COVID-19 Vaccine Efficacy Trial. medRxiv (2021).

41. Earle, K.A. et al. Evidence for antibody as a protective correlate for COVID-19 vaccines. Vaccine 39, 4423–4428 (2021).

42. Khoury, D.S. et al. Neutralizing antibody levels are highly predictive of immune protection from symptomatic SARS-CoV-2 infection. Nature Medicine 27, 1205–1211 (2021).

43. Voysey, M. et al. Safety and efficacy of the ChAdOx1 nCoV-19 vaccine (AZD1222) against SARS-CoV-2: an interim analysis of four randomised controlled trials in Brazil, South Africa, and the UK. Lancet 397, 99–111 (2021).

44. Wood, S.N. Generalized additive models: an introduction with R. chapman and hall/CRC, 2006.

45. Cho, A. et al. Anti-SARS-CoV-2 receptor-binding domain antibody evolution after mRNA vaccination. Nature 2021 600:7889 600, 517-522 (2021).

46. Kim, P., Gordon, S.M., Sheehan, M.M. & Rothberg, M.B. Duration of SARS-CoV-2 Natural Immunity and Protection against the Delta Variant: A Retrospective Cohort Study. Clinical Infectious Diseases (2021).

47. Goldberg, Y., et al. Protection and waning of natural and hybrid COVID-19 immunity. medRxiv, 2021.2012.2004.21267114-21262021.21267112.21267104.21267114 (2021).

48. Feikin, D., et al. Duration of Effectiveness of Vaccines Against SARS-CoV-2 Infection and COVID-19 Disease: Results of a Systematic Review and Meta-Regression. SSRN Electronic Journal (2021).

49. Papa, G. et al. Furin cleavage of SARS-CoV-2 Spike promotes but is not essential for infection and cell-cell fusion. PLoS pathogens 17, e1009246–e1009246 (2021).

50. Braga, L. et al. Drugs that inhibit TMEM16 proteins block SARS-CoV-2 Spike-induced syncytia. Nature 594, 88–93 (2021).

51. Zhang, J. et al. Membrane fusion and immune evasion by the spike protein of SARS-CoV-2 Delta variant. Science, eabl9463–eabl9463 (2021).

52. Kodaka, M. et al. A new cell-based assay to evaluate myogenesis in mouse myoblast C2C12 cells. Experimental cell research 336, 171–181 (2015).

53. Abdelnabi, R., et al. The omicron (B.1.1.529) SARS-CoV-2 variant of concern does not readily infect Syrian hamsters. bioRxiv, 2021.2012.2024.474086-472021.474012.474024.474086 (2021).

54. Jackson, C.B., Farzan, M., Chen, B. & Choe, H. Mechanisms of SARS-CoV-2 entry into cells. Nature Reviews Molecular Cell Biology 23, 3–20 (2022).

55. Mykytyn, A.Z. et al. SARS-CoV-2 entry into human airway organoids is serine protease-mediated and facilitated by the multibasic cleavage site. Elife 10, e64508–e64508 (2021).

56. Hoffmann, M. et al. SARS-CoV-2 cell entry depends on ACE2 and TMPRSS2 and is blocked by a clinically proven protease inhibitor. cell 181, 271–280 (2020).

57. Hoffmann, M., Kleine-Weber, H. & Pöhlmann, S. A multibasic cleavage site in the spike protein of SARS-CoV-2 is essential for infection of human lung cells. Molecular cell 78, 779–784 (2020).

58. Nie, J. et al. Functional comparison of SARS-CoV-2 with closely related pangolin and bat coronaviruses. Cell discovery 7, 1–12 (2021).

59. Wrobel, A.G. et al. Structure and binding properties of Pangolin-CoV spike glycoprotein inform the evolution of SARS-CoV-2. Nat. Commun. 12, 1–6 (2021).

60. Belouzard, S., Chu, V.C. & Whittaker, G.R. Activation of the SARS coronavirus spike protein via sequential proteolytic cleavage at two distinct sites. Proceedings of the National Academy of Sciences 106, 5871–5876 (2009).

61. Dicken, S.J., et al. Characterisation of B. 1.1. 7 and Pangolin coronavirus spike provides insights on the evolutionary trajectory of SARS-CoV-2. bioRxiv (2021).

62. Saccon, E. et al. Cell-type-resolved quantitative proteomics map of interferon response against SARS-CoV-2. Iscience 24, 102420–102420 (2021).

63. Winstone, H. et al. The polybasic cleavage site in SARS-CoV-2 spike modulates viral sensitivity to type I interferon and IFITM2. Journal of virology 95, e02422–02420 (2021).

64. Daniloski, Z. et al. Identification of required host factors for SARS-CoV-2 infection in human cells. Cell 184, 92–105 (2021).

65. Sheikh, A., Kerr, S., Woolhouse, M., McMenamin, J. & Robertson, C. Severity of omicron variant of concern and vaccine effectiveness against symptomatic disease: national cohort with nested test negative design study in Scotland. (2021).

66. Report 50 - Hospitalisation risk for Omicron cases in England | Faculty of Medicine | Imperial College London.

67. Benvenuto, D. et al. Evolutionary analysis of SARS-CoV-2: how mutation of Non-Structural Protein 6 (NSP6) could affect viral autophagy. Journal of Infection 81, e24–e27 (2020).

68. Thorne, L.G. et al. Evolution of enhanced innate immune evasion by SARS-CoV-2. Nature, 1–12 (2021).

69. Rihn, S.J. et al. A plasmid DNA-launched SARS-CoV-2 reverse genetics system and coronavirus toolkit for COVID-19 research. PLoS biology 19, e3001091–e3001091 (2021).

70. Davis, C. et al. Reduced neutralisation of the Delta (B. 1.617. 2) SARS-CoV-2 variant of concern following vaccination. PLoS pathogens 17, e1010022–e1010022 (2021).

71. Hughes, E.C. et al. SARS-CoV-2 serosurveillance in a patient population reveals differences in virus exposure and antibody-mediated immunity according to host demography and healthcare setting. J Infect Dis (2020).

72. Newman, J., et al. Neutralising antibody activity against SARS-CoV-2 variants, including Omicron, in an elderly cohort vaccinated with BNT162b2. medRxiv, 2021.2012.2023.21268293-21262021.21268212.21268223.21268293 (2021).

73. Zufferey, R., Nagy, D., Mandel, R.J., Naldini, L. & Trono, D. Multiply attenuated lentiviral vector achieves efficient gene delivery in vivo. Nat.Biotechnol. 15, 871–875 (1997).

74. Zufferey, R. et al. Self-inactivating lentivirus vector for safe and efficient in vivo gene delivery. J Virol 72, 9873–9880 (1998).

75. Li, H. Minimap2: pairwise alignment for nucleotide sequences. Bioinformatics 34, 3094–3100 (2018).

76. Price, M.N., Dehal, P.S. & Arkin, A.P. FastTree 2–approximately maximum-likelihood trees for large alignments. PloS one 5, e9490–e9490 (2010).

77. Woo, H. et al. Developing a fully glycosylated full-length SARS-CoV-2 spike protein model in a viral membrane. The Journal of Physical Chemistry B 124, 7128–7137 (2020).

78. Liu, Z. et al. Identification of SARS-CoV-2 spike mutations that attenuate monoclonal and serum antibody neutralization. Cell host & microbe 29, 477–488 (2021).

79. Lan, J. et al. Structure of the SARS-CoV-2 spike receptor-binding domain bound to the ACE2 receptor. Nature 581, 215–220 (2020).

80. Pymol, T. The PyMOL molecular graphics system. *Version* 1, r3pre-r3pre (2010).

